# Differences in Cardiac Mechanics among Genetically At-Risk First-Degree Relatives: The DCM Precision Medicine Study

**DOI:** 10.1101/2023.05.30.23290123

**Authors:** Jane E. Wilcox, Lauren Beussink-Nelson, Jinwen Cao, Ritika Kumar, Elizabeth Jordan, Hanyu Ni, Sanjiv J. Shah, Ray E. Hershberger, Daniel D. Kinnamon

**Affiliations:** Division of Cardiology, Department of Medicine, Northwestern University Feinberg School of Medicine, Chicago, IL; Division of Human Genetics, Department of Internal Medicine, The Ohio State University, Columbus, OH; The Davis Heart and Lung Research Institute, The Ohio State University, Columbus, OH; Division of Cardiovascular Medicine, Department of Internal Medicine, The Ohio State University, Columbus, OH

**Keywords:** cardiomyopathy, echocardiography, genetics, family-based study

## Abstract

**Aims:** Among genetically at-risk first-degree relatives (FDRs) of probands with dilated cardiomyopathy (DCM), the ability to detect changes in left ventricular (LV) mechanics with normal LV size and ejection fraction (LVEF) remains incompletely explored. We sought to define a pre-DCM phenotype among at-risk FDRs, including those with variants of uncertain significance (VUSs), using echocardiographic measures of cardiac mechanics.

**Methods and Results:** LV structure and function, including speckle-tracking analysis for LV global longitudinal strain (GLS), were evaluated in 124 FDRs (65% female; median age 44.9 [IQR: 30.6-60.3] years) of 66 DCM probands of European ancestry sequenced for rare variants in 35 DCM genes. FDRs had normal LV size and LVEF. Negative FDRs of probands with pathogenic or likely pathogenic (P/LP) variants (n=28) were a reference group to which negative FDRs of probands without P/LP variants (n=30), FDRs with only VUSs (n=27), and FDRs with P/LP variants (n=39) were compared. In an analysis accounting for age-dependent penetrance, FDRs below the median age showed minimal differences in LV GLS across groups while those above it with P/LP variants or VUSs had lower absolute values than the reference group (−3.9 [95% CI: −5.7, −2.1] or −3.1 [−4.8, −1.4] %-units) and negative FDRs of probands without P/LP variants (−2.6 [−4.0, −1.2] or −1.8 [−3.1, −0.6]).

**Conclusions:** Older FDRs with normal LV size and LVEF who harbored P/LP variants or VUSs had lower absolute LV GLS values, indicating that some DCM-related VUSs are clinically relevant. LV GLS may have utility for defining a pre-DCM phenotype.

**Clinical Trial Registration:** clinicaltrials.gov, NCT03037632

**Graphical Abstract:** **Differences in Cardiac Mechanics among Genetically At-Risk First-Degree Relatives: The DCM Precision Medicine Study.** DCM = dilated cardiomyopathy, FDR = first-degree relative, GLS = global longitudinal strain, HF = heart failure, LP = likely pathogenic, LV = left ventricular, LVEF = left ventricular ejection fraction, P = pathogenic, VUS = variant of uncertain significance.

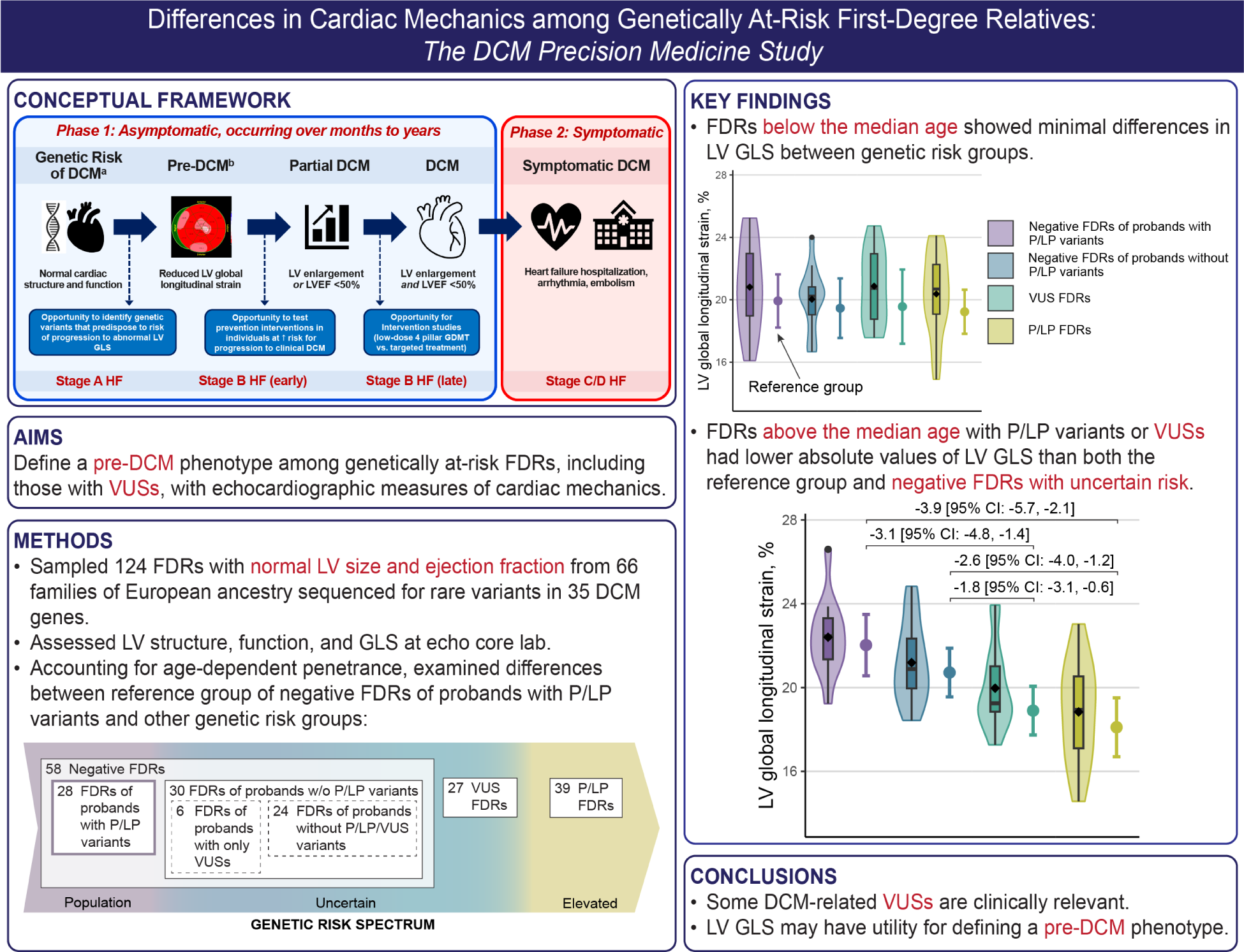

## INTRODUCTION

It is now well established that idiopathic dilated cardiomyopathy (DCM) has a genetic basis, and that genetic analysis to identify variants in relevant DCM genes can be used to assess genetic risk in patients (probands) and their asymptomatic first-degree relatives (FDRs).^1,2^ Cascade genetic testing of FDRs (parents, full siblings, and children) for pathogenic (P) or likely pathogenic (LP) variants identified in a DCM proband is a well-established practice to assess FDR genetic risk, and can be used for predictive testing and to tailor management recommendations.^3^ Those FDRs testing positive for P/LP variants who do not yet have DCM are recommended to have ongoing clinical surveillance for evidence of emergent DCM. Conversely, FDRs who test negative are considered to be at population risk or lower and can be released from surveillance, as a P/LP variant is considered a primary cause of DCM using current variant interpretation guidance.^3–6^

The DCM Precision Medicine Study, a family-based study of DCM probands and their relatives,^7^ has also contributed data in support of a genetic basis of DCM.^6,8^ Probands in the DCM Precision Medicine Study,^6–8^ the parent study of the current investigation, underwent genetic analysis to identify variants classified as P, LP, or of uncertain significance (VUS) in relevant DCM genes, with 15% of probands having P/LP variants, 46% having only VUSs, and 38% having no P/LP/VUS variant identified in an initial sample.^6^ Enrolled relatives were also sequenced for P/LP/VUS variants identified in their proband to assess their level of genetic risk. For FDRs with only VUSs, genetic risk remained uncertain because a VUS shared with the proband may or may not be disease-associated. Also, in probands for whom no P/LP/VUS variants were identified, unmeasured genetic risk factors for DCM could still be present and shared with FDRs.^9^

VUSs are not recommended to be used for predictive testing due to the inherent uncertainty of their relationship to DCM. Defining this relationship, if any, to the DCM phenotype in a patient or family is highly clinically relevant given the preponderance of probands with only VUSs.^6^ Nonetheless, a clinical approach to evaluate the impact of VUSs has not been reported. Prior work using speckle-tracking echocardiography (STE) found that subtle abnormalities in global longitudinal strain (GLS) were present in FDRs from five families who carried P/LP variants in *MYH7*, *TPM1*, or *TNNT2* despite having normal LV ejection fraction (LVEF) and size.^10^ Additional studies have validated the usefulness of STE to identify LV strain abnormalities in FDRs of DCM probands,^11–14^ as recently reviewed.^15^ However, cardiac mechanics have not been investigated by STE in FDRs who harbor only VUSs.

An ancillary study of the DCM Precision Medicine Study was established to analyze echocardiographic data to test the hypotheses that (1) DCM-related abnormalities in cardiac mechanics were present in FDRs genetically at risk for DCM before development of LV systolic dysfunction (LVSD) and dilation, and (2) that differences in cardiac mechanics would reflect the level of genetic risk (Figure 1). In this report, echocardiographic measurements were compared in FDRs with normal LV size and LVEF grouped by age and level of genetic risk (P/LP, VUS, or negative), including a reference group of FDRs of probands with P/LP variants who tested negative for their proband’s variants and should have DCM risk no greater than the general population.

**Figure 1.**
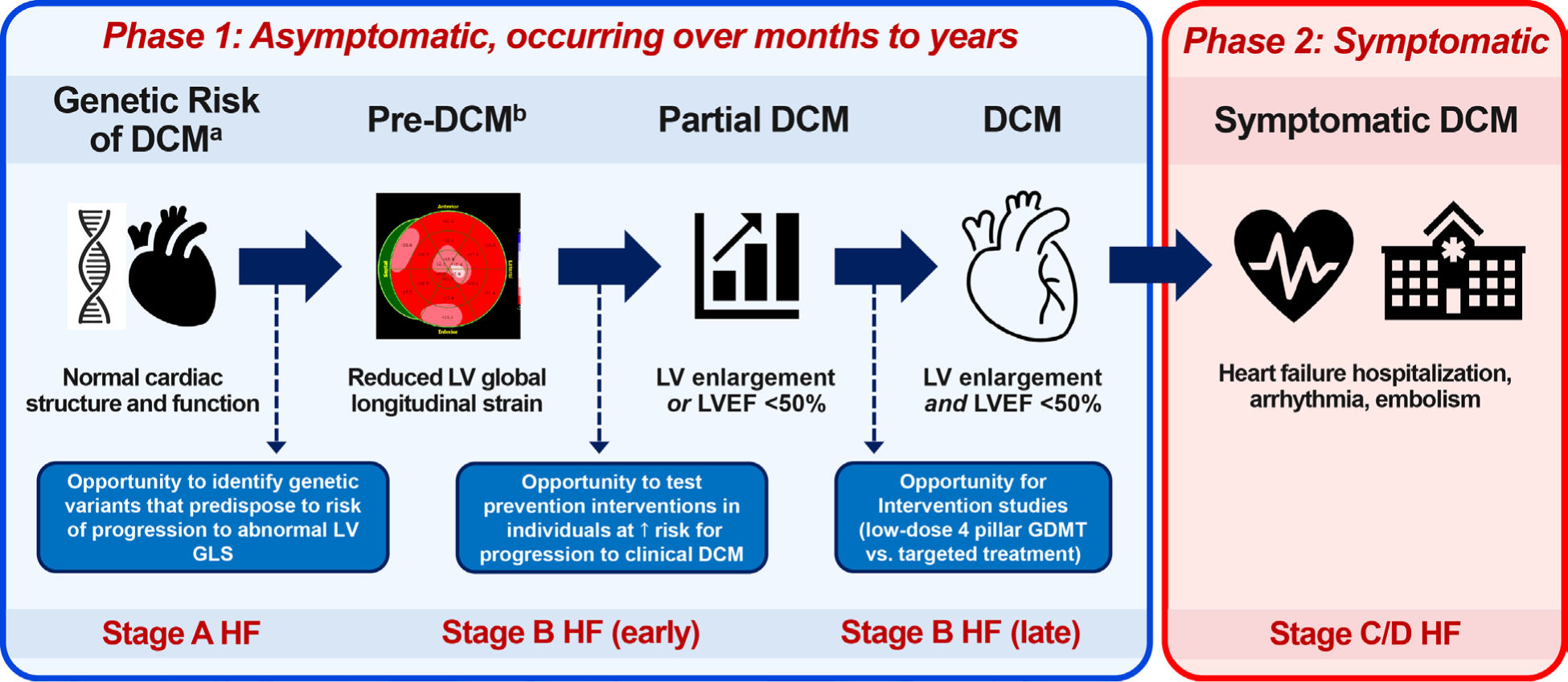
A conceptual framework for a pre-DCM phenotype. Identifying pre-DCM in at-risk individuals using LV GLS provides an opportunity to test interventions to prevent DCM at an earlier stage. Pre-DCM^a^ is defined as a reduction in the absolute value of LV GLS in a genetically at-risk first-degree relative who still has normal LV size and ejection fraction. Partial DCM^b^ has been defined as an LVEF <50% or LV enlargement (but not both),^8,30^ and has been shown to be a harbinger of DCM. The transition from genetic risk of DCM with normal cardiac structure and function to pre-DCM, partial DCM, clinically identifiable though still asymptomatic DCM (Phase 1), and finally symptomatic DCM (Phase 2) takes months or years. ACC/AHA stages of heart failure aligned with the progression of DCM are shown at the bottom. HF = heart failure.

## METHODS

### Participants and informed consent

The DCM Precision Medicine Study is a multi-site consortium-based cross-sectional study of families.^7^ Accrual of DCM probands and family members occurred from June 7, 2016 to March 15, 2020 (probands) and April 1, 2021 (family members). The investigation conforms with the principles outlined in the *Declaration of Helsinki*. The Institutional Review Boards (IRBs) at The Ohio State University (OSU) and all clinical sites approved the initial period of the study followed by oversight by a single IRB at the University of Pennsylvania. Written informed consent was obtained from all participants.

### Demographic and clinical data collection

At the time of enrollment, study personnel collected a cardiovascular history and a pedigree in a standardized interview. Structured interviews collected participant social demographics (e.g., age at enrollment, sex, years of education, tobacco use) and self-reported medical history; medical record questionnaires validated and summarized any prior key cardiovascular clinical information that was available. FDRs underwent clinical screening with two-dimensional trans-thoracic echocardiograms, either as part of the parent study or with their clinician.

The DCM Consortium is aware of issues regarding the collection, analysis, presentation and discussion of race, ethnicity and ancestry and has adopted recommended approaches.^16^ Because FDR genetic risk groups defined below were based on the proband’s variants shared by the FDR, proband genomic ancestry, rather than self-reported race and ethnicity in the proband or FDR, was used for adjustment. Proband global ancestry proportions were inferred from Illumina Global Screening Array genotypes with the 1000 Genomes Phase 3 integrated call set as the reference (Supplemental Methods).

### Echocardiographic data acquisition and analysis

Clinical and study echocardiograms were digitized and sent to OSU for processing and storage. An in-house pipeline leveraging DICOM Toolkit v3.6.6 (OFFIS; https://dcmtk.org) and DicomAnonymizerTool (Radiological Society of North America; https://mircwiki.rsna.org) was built to re-encode images to a standard format and remove personally identifiable information. The resulting images were reviewed by two individuals to ensure removal of all identifiers and then transferred electronically to the Northwestern University Echocardiography Core Laboratory (NUECL) for analysis.

Echocardiographic images were centrally quantified at the NUECL by two experienced readers (LN, RG) with expertise in conventional echocardiography and STE analysis. Readers were blinded to all other study participant data, including variant status. Digitized cine loops were analyzed using TomTec-Arena software version TTA 2.50 (Tomtec Imaging Systems). Image quality scores were assigned based on the percentage of the endocardium visualized (1=0%-25%, 2=26%-50%, 3=51%-75%, 4=76%-100%). Standard measurements of LV structure and function were made according to criteria established by the American Society of Echocardiography.^17^ In brief, LV dimensions and wall thickness were measured from the parasternal long-axis view perpendicular to the long axis of the LV at end diastole and end systole. LV volumes were measured using the method of disks in the apical 4- and 2-chamber views at end diastole and end systole. Volumes were indexed by dividing by Mosteller body surface area.^18^ LVEF was quantified using the modified Simpson’s method based on the LV volumes measured in the apical 4- and 2-chamber views. Single-plane LV volumes and LVEF were recorded if volumes were not quantifiable in both planes.

LV GLS was quantified by tracing the endocardial border at end systole in the apical 4-, 2-, and 3-chamber views using a semi-automated package within the TomTec-Arena software (LV AutoStrain). If two or more continuous segments were not well visualized, images were significantly foreshortened, or images were acquired off-axis, speckle tracking was not attempted. If tracking of wall motion did not appear adequate based on visual inspection, the region of interest was manually adjusted to improve the quality of the tracking. Components of LV strain were recorded as previously described.^19^ Because LV GLS was not quantifiable in all subjects, LV longitudinal strain in the apical 4-chamber view (LSA4C), which was available for all subjects, was also analyzed. For ease of reporting and interpretation, all strain values were reported as absolute values (lower absolute strain values correspond to worse cardiac mechanics).

### Genetic Analysis

Research exome sequencing of DCM probands was conducted at the University of Washington Northwest Genomics Center, and genomic data files were transferred to the Division of Human Genetics Data Management Platform at the Ohio Supercomputer Center for analysis of a panel of 35 genes considered clinically relevant for DCM.^6^ Variants were adjudicated using American College of Medical Genetics (ACMG)^4^ and ClinGen-based criteria tailored to DCM^6^ and assigned to an ACMG category (Supplemental Methods). P, LP, and VUS variants were confirmed in the proband and cascade-tested in FDRs by Sanger sequencing.

### Sample Selection

FDRs were selected from a sampling frame of 788 with no evidence of LVSD or LV enlargement (LVE) per parent study adjudication, available images from the study diagnostic echo, and cascade Sanger genotyping for all P/LP/VUS variants found in their proband as of October 20, 2021 (Figure 2). To ensure adequate representation in each genetic risk group, approximately 50 FDRs were selected from each of three strata defined by the most deleterious variant harbored by the FDR (none, VUS, or P/LP), with preference given to unrelated FDRs. To expand the sample, all remaining FDRs in the sampling frame belonging to families of FDRs selected in the initial round were added without regard to variants harbored, yielding data for 244 FDRs from 124 families. Because DCM is an adult-onset disease with age-dependent penetrance, FDRs <18 years of age (n=29), who are less likely to have a detectable phenotype, were excluded. Remaining FDRs with non-interpretable images (n=5), LVSD (LVEF <50%) or LVE (end-diastolic internal diameter ≥95^th^ percentile for sex and height^20^) upon analysis at NUECL (n=17), or missing proband ancestry (n=1) were also excluded. Proband ancestry varied substantially across genetic risk groups defined below, but the reference group included only 2 FDRs of probands with less than 87.5% European ancestry, each with markedly different proband non-European ancestry proportions. Because comparisons of genetic risk groups could not be reliably adjusted for proband ancestry in the full sample with such sparse data, we restricted analysis to FDRs of probands with 87.5% or more European ancestry, who were well represented in all genetic risk groups. The final data set comprised echocardiograms of 124 FDRs from 66 families.

**Figure 2.**
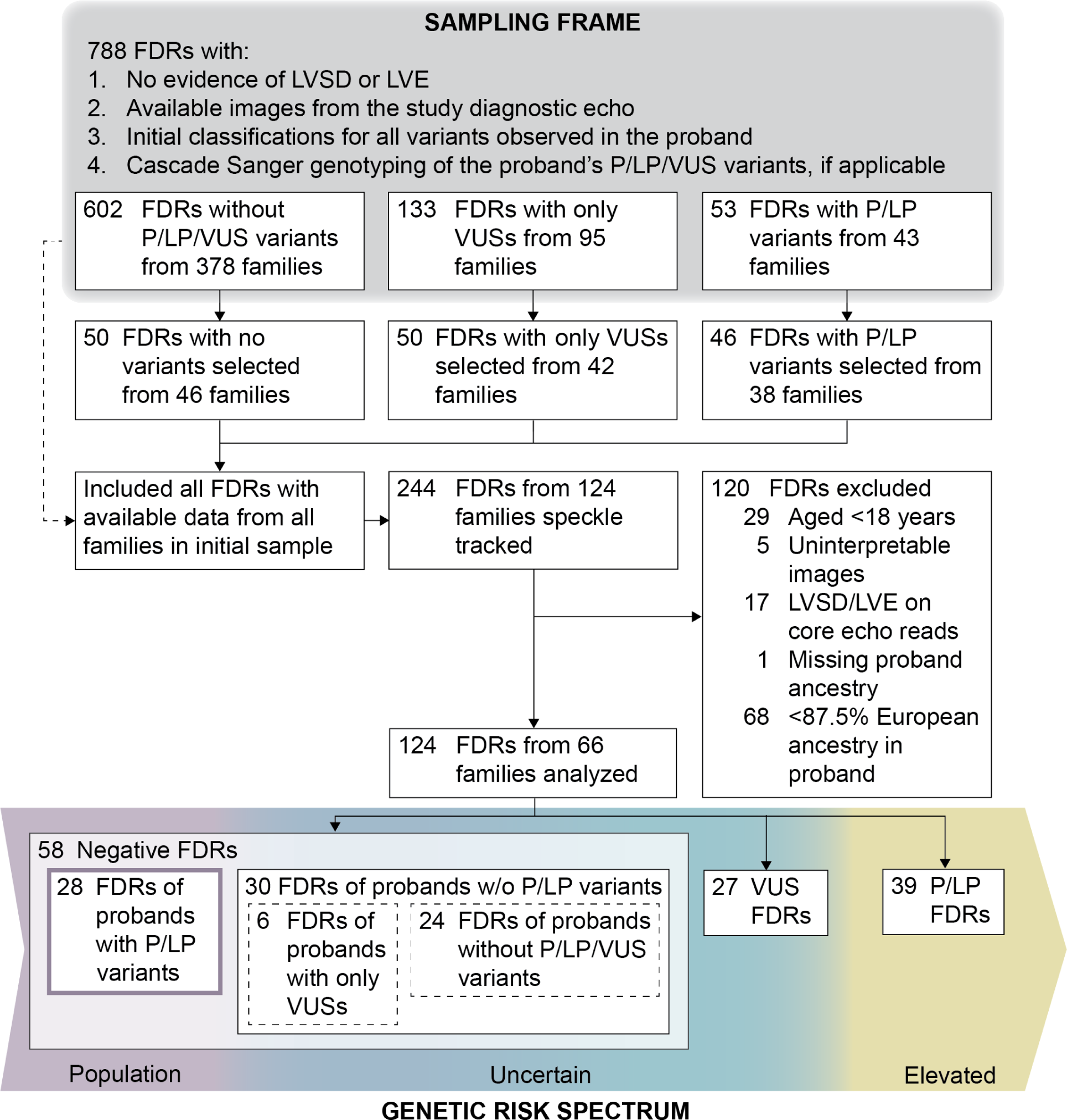
Selection of study participants and definition of genetic risk groups. FDRs of probands who had completed genetic analysis were selected from a sampling frame including FDRs meeting all specified criteria as of October 20, 2021. To ensure adequate representation in each genetic risk group, approximately 50 FDRs were selected from each of three strata defined by the most deleterious variant harbored by the FDR (none, VUS, or P/LP), with preference given to unrelated FDRs. To expand the sample, all remaining FDRs in the sampling frame belonging to families of FDRs selected in the initial round were added without regard to variants harbored. For analysis, FDRs <18 years of age were excluded because DCM is an adult-onset disease with age-dependent penetrance, as were FDRs of probands with less than 87.5% European ancestry, who were not represented in sufficient numbers in the reference group described below to permit statistical adjustment of group comparisons by ancestry in the full sample. FDRs were analyzed in groups with varying levels of genetic risk, with negative FDRs of probands with P/LP variants serving as the reference group with DCM risk no greater than the general population.

### Genetic risk groups

FDRs were assigned to one of three genetic risk groups based on the most deleterious of the proband’s variants harbored, with FDRs who did not harbor any P/LP/VUS variants further divided into two subgroups depending upon their proband’s genetics (Figure 2).

1. **P/LP FDRs** (n=39) included those who harbored any of the proband’s P/LP variants and were therefore at elevated genetic risk.
2. **VUS FDRs** (n=27) included those who harbored only VUS variants found in the proband and therefore had uncertain genetic risk. Note that probands of VUS FDRs may have also carried a P/LP variant that was not shared with the VUS FDRs.
3. **Negative FDRs** (n=58) included those with no P/LP/VUS variants identified and were subdivided into two groups because their genetic risk was dependent upon the findings in the proband:

a. **Negative FDRs of probands with P/LP variants (reference)** (n=28) are generally released from surveillance for having risk no greater than the general population and were therefore used as the reference group. Note that probands of FDRs in this group also may have had VUSs in addition to their P/LP variant that were not shared with their FDRs.
b. **Negative FDRs of probands without P/LP variants** (n=30), of which 6 were FDRs of probands with only VUSs and 24 were FDRs of probands with no P/LP/VUS variants identified, had uncertain genetic risk because a negative genetic test result did not eliminate a yet unknown genetic cause of DCM in the family unit. While unmeasured genetic risk may be lower for negative FDRs of probands with only VUSs, the small number of FDRs in this subgroup precluded further subdivision, and all negative FDRs of probands without P/LP variants were analyzed as a single group.

### Statistical analysis

All analyses were performed in R version 4.0.2 (R Foundation) and SAS/STAT 15.2 software, Version 9.4 (TS1M7) of the SAS System for 64-bit Windows (SAS Institute). Under a threshold model of DCM development that explains both age-dependent penetrance and variable expressivity, quantitative measures of cardiac structure and function that are similar at younger ages worsen more rapidly with age in individuals with higher genetic risk, leading to differences between genetic risk groups that start small and grow with age.^9^ To reflect this growth without assuming linear age trajectories, the mean of each echocardiographic measurement was modeled as a function of genetic risk group within two age groups (below or above the sample median age at echocardiogram) using a single linear mixed model with an interaction.^21^ For all measurements other than sex-specific internal diameter z-scores, sex and its interaction with age group were included as covariates. For LV GLS and LSA4C, height, weight, and image quality rating (≤2 vs. >2) were also included. Heterogeneity between clinical sites and intrafamilial correlation were modeled by including independent normal random effects for proband enrollment site and family within site.

This model was fit with restricted maximum likelihood using SAS/STAT PROC GLIMMIX, and the Morel-Bokossa-Neerchal bias-corrected empirical covariance matrix with sites as independent units and the standard normal distribution were used for inference on the fixed effects.^21^ This approach should have yielded asymptotically valid inferences on the fixed effects if only the mean was correctly specified,^22^ which was desirable because the compound symmetric within-family working covariance matrix implied by the model was an approximation. All statistical tests and confidence intervals were two-sided with significance level of 0.05.

## RESULTS

The median age of FDRs was 44.9 years (IQR: 30.6–60.3), and 65% were female. Clinical characteristics of the 124 FDRs analyzed are shown by genetic risk group, as defined above (Table 1; Supplemental Table 1). Comorbid conditions had prevalence comparable to or lower than the general population, with 36% classified as obese, 15% having reported history of hypertension, 5% with hyperlipidemia and 2% having diabetes. Because FDRs included in this study could not have evidence of LVSD or LVE, all usual echocardiographic clinical measures were within normal limits (Table 2; Supplemental Table 2).

**Table 1.** Demographic characteristics and comorbidities of first-degree relatives, by genetic risk group

| Characteristic | Negative FDRs<br>(N = 58) |  | VUS FDRs<br>(N = 27) | P/LP FDRs<br>(N = 39) |
| --- | --- | --- | --- | --- |
|  | *FDRs of probands with P/LP variants (reference)*<br>(N = 28) | *FDRs of probands without P/LP variants*<br>(N = 30) |  |  |
| **Demographics** |  |  |  |  |
| Age at echocardiogram, years, median (IQR) | 43.6 (34.3, 60.2) | 52.8 (39.5, 62.7) | 45.7 (28.7, 57.2) | 39.3 (28.2, 59.4) |
| Age at echocardiogram at or above median (44.9 years), No. (%) | 14 (50.0) | 16 (53.3) | 14 (51.9) | 18 (46.2) |
| Female, No. (%) | 18 (64.3) | 19 (63.3) | 17 (63.0) | 27 (69.2) |
| Proband ancestry, %, median (min - max) |  |  |  |  |
| European | 97.3 (94.7, 99.1) | 96.8 (94.7, 99.0) | 96.2 (89.6, 98.7) | 96.9 (89.8, 99.1) |
| African | 0.4 (0.0, 1.3) | 0.2 (0.0, 1.7) | 0.4 (0.0, 1.9) | 0.5 (0.0, 2.6) |
| Native American | 0.2 (0.0, 1.8) | 0.6 (0.1, 1.4) | 0.7 (0.0, 7.3) | 0.5 (0.0, 4.8) |
| Other | 1.6 (0.0, 4.4) | 2.0 (0.6, 4.4) | 2.4 (0.5, 5.2) | 2.0 (0.0, 5.5) |
| **Comorbidities, No. (%)** |  |  |  |  |
| Obesity (BMI $\geq 30$ kg/m<sup>2</sup>) | 15 (53.6) | 14 (46.7) | 6 (22.2) | 10 (25.6) |
| Hypertension | 4 (14.3) | 6 (20.0) | 3 (11.1) | 5 (12.8) |
|  | *FDRs of probands with P/LP variants (reference)*<br>(N = 28) | *FDRs of probands without P/LP variants*<br>(N = 30) |  |  |
| High Cholesterol | 2 (7.1) | 1 (3.3) | 0 (0.0) | 3 (7.7) |
| Diabetes | 0 (0.0) | 1 (3.3) | 1 (3.7) | 0 (0.0) |
| Tobacco use (ever), No. (%) | 5 (17.9) | 7 (23.3) | 8 (29.6) | 8 (20.5) |
| Years smoked, mean (SD) | 23.0 (11.0)<br>[n=5] | 12.4 (6.2)<br>[n=7] | 21.7 (12.9)<br>[n=7] | 10.5 (10.7)<br>[n=8] |
| Cigarettes per day, mean (SD) | 20.3 (10.3)<br>[n=4] | 14.0 (13.9)<br>[n=7] | 12.7 (7.9)<br>[n=7] | 11.4 (8.5)<br>[n=5] |
| Alcohol use (ever) and consume $\geq 5$ drinks in an occasion, No. (%) | 2 (7.1) | 2 (6.7) | 2 (7.4) | 4 (10.3) |
| Alcohol use frequency, No. (%) | [n=25] | [n=17] | [n=16] | [n=35] |
| 1-3 times/month or <1 time/month | 8 (32.0) | 10 (58.8) | 11 (68.8) | 6 (17.1) |
| 1-3 times/week | 14 (56.0) | 4 (23.5) | 4 (25.0) | 16 (45.7) |
| 4-7 times/week | 3 (12.0) | 3 (17.6) | 1 (6.3) | 13 (37.1) |

Abbreviations: FDR = first-degree relative; P = pathogenic; LP = Likely pathogenic; VUS = variant of uncertain significance.

**Table 2.** Echocardiographic measurements of first-degree relatives, by genetic risk group

| **Echocardiographic Measurement** | **Negative FDRs**<br>**(n = 58)** |  | **VUS FDRs**<br>**(N = 27)** | **P/LP FDRs**<br>**(N = 39)** |
| --- | --- | --- | --- | --- |
|  | *FDRs of probands with P/LP variants (reference)*<br>*(N = 28)* | *FDRs of probands without P/LP variants*<br>*(N = 30)* |  |  |
| LV global longitudinal strain<sup>a</sup>, %, mean (SD) | 21.5 (2.5)<br>[n=26] | 20.7 (2.0) | 20.4 (2.2)<br>[n=25] | 19.6 (2.6)<br>[n=35] |
| LV global longitudinal strain category<sup>a</sup>, No. (%) | [n=26] |  | [n=25] | [n=35] |
| ≥18% (normal) | 23 (88.5) | 27 (90.0) | 23 (92.0) | 26 (74.3) |
| ≥16%, <18% (borderline) | 3 (11.5) | 3 (10.0) | 2 (8.0) | 6 (17.1) |
| <16% (abnormal) | 0 (0.0) | 0 (0.0) | 0 (0.0) | 3 (8.6) |
| LV longitudinal strain (A4C view), %, mean (SD) | 21.5 (2.8) | 20.2 (2.6) | 20.7 (2.4) | 19.8 (3.1) |
| LV ejection fraction, %, mean (SD) | 61.8 (3.9) | 61.0 (3.6) | 60.9 (3.6) | 59.5 (3.5) |
| LV internal diameter at end diastole, z-score<sup>b</sup>, mean (SD) | -0.6 (1.5) | -0.1 (1.3) | -1.1 (1.9) | -0.3 (1.1)<br>[n=38] |
| LV internal diameter at end systole, z-score<sup>b</sup>, mean (SD) | 0.1 (1.2) | 0.7 (1.5) | 0.0 (1.1) | 0.8 (1.2)<br>[n=38] |
| Indexed LV end diastolic volume<sup>c</sup>, ml/m<sup>2</sup>, mean (SD) | 53.0 (9.5) | 50.4 (10.2) | 50.2 (10.7) | 51.6 (10.2) |
| Indexed LV end systolic volume<sup>c</sup>, ml/m<sup>2</sup>, mean (SD) | 20.3 (4.3) | 19.7 (4.6) | 19.6 (4.4) | 20.9 (4.7) |
|  | *FDRs of probands with P/LP variants (reference)*<br>(N = 28) | *FDRs of probands without P/LP variants*<br>(N = 30) |  |  |
| Posterior wall thickness at end diastole, mm, mean (SD) | 8.1 (1.3) | 8.6 (1.4) | 7.9 (1.2) | 8.1 (1.3)<br>[n=38] |
| Septal wall thickness at end diastole, mm, mean (SD) | 9.8 (2.5) | 9.4 (1.7) | 8.8 (1.8) | 8.7 (2.2)<br>[n=38] |

Abbreviations: A4C = apical 4-chamber; FDR = first-degree relative; P = pathogenic; LP = Likely pathogenic; LV = left ventricular; VUS = variant of uncertain significance.
<sup>a</sup> LV global longitudinal strain could not be quantified in n = 8 FDRs who were missing one or more of the apical 2- or 3-chamber views.
<sup>b</sup> Calculated based on sex and height<sup>20</sup> for all study participants with heights of at least 152 cm (male) or 137 cm (female).
<sup>c</sup> Calculated using volume divided by Mosteller body surface area.

Based on a model accounting for the age-dependent emergence of DCM, differences in quantitative measures of cardiac structure and function between genetic risk groups were expected to start small and grow with age, so a single linear mixed model was used to estimate these differences within two age groups (below or above the sample median age at echocardiogram). Among FDRs below the median age, differences between other genetic risk groups and the reference group (negative FDRs of probands with P/LP variants) were typically small (Table 3; Supplemental Figure 1). Among FDRs above the median age in the sample, larger differences between genetic risk groups were observed for LV GLS (Table 3; Supplemental Table 3; Figure 3). In particular, P/LP FDRs in this age group, who had elevated genetic risk, had lower absolute LV GLS values than the reference group with population risk and negative FDRs of probands without P/LP variants, who had uncertain risk. VUS FDRs in this age group, who had uncertain genetic risk, also had lower absolute LV GLS values than these two groups. These findings were confirmed using LV LSA4C measurements available for all subjects. While point estimates also suggested that P/LP and VUS FDRs had lower LVEF and septal wall thickness relative to the reference group, confidence intervals did not rule out the possibility of no difference.

**Figure 3.**
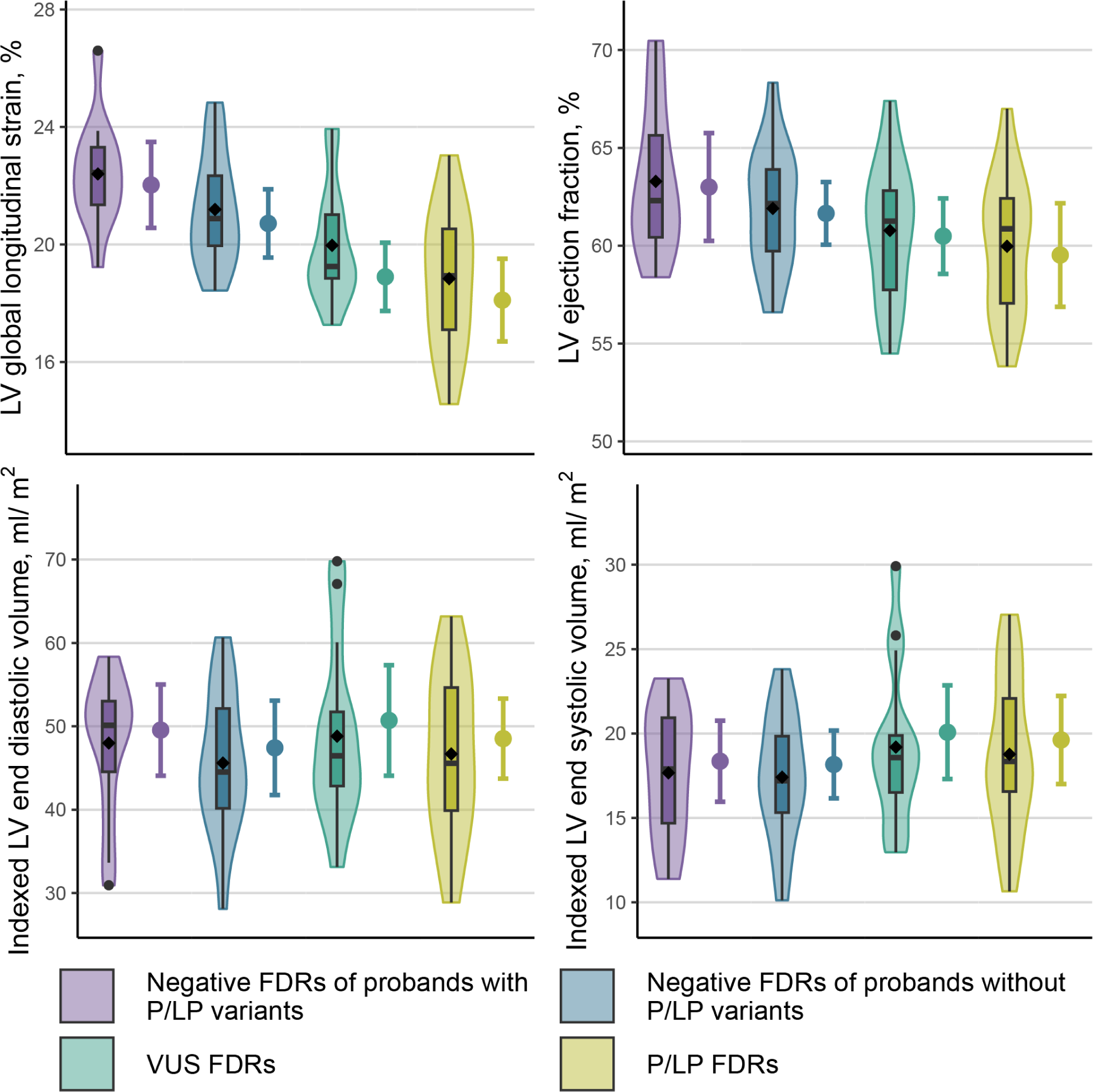
Echocardiographic measurements by genetic risk group in FDRs above the median age in the sample. A violin plot with a superimposed box-and-whisker plot shows the distribution of the measurements among FDRs in each genetic risk group, with a black diamond at the mean. Next to this, an interval plot shows the estimated marginal mean from the linear mixed model analysis (point) as well as its 95% confidence interval (interval) obtained using Morel-Bokossa-Neerchal bias-corrected empirical standard errors and the standard normal distribution. For each genetic risk group, the estimated marginal mean is a covariate-adjusted estimate of the mean in a population of FDRs above the median age in the sample (44.9 years) that is half female. For LV global longitudinal strain, these populations also have the same mean height and weight and half image quality >2. Table 3 presents covariate-adjusted estimated mean differences between each genetic risk group and the reference group for FDRs above the median age from the same model; Supplemental Table 3 presents these differences comparing the P/LP and VUS groups to negative FDRs of probands without P/LP variants. In both tables, reported differences are the differences between the estimated marginal means in this figure.

**Table 3.**
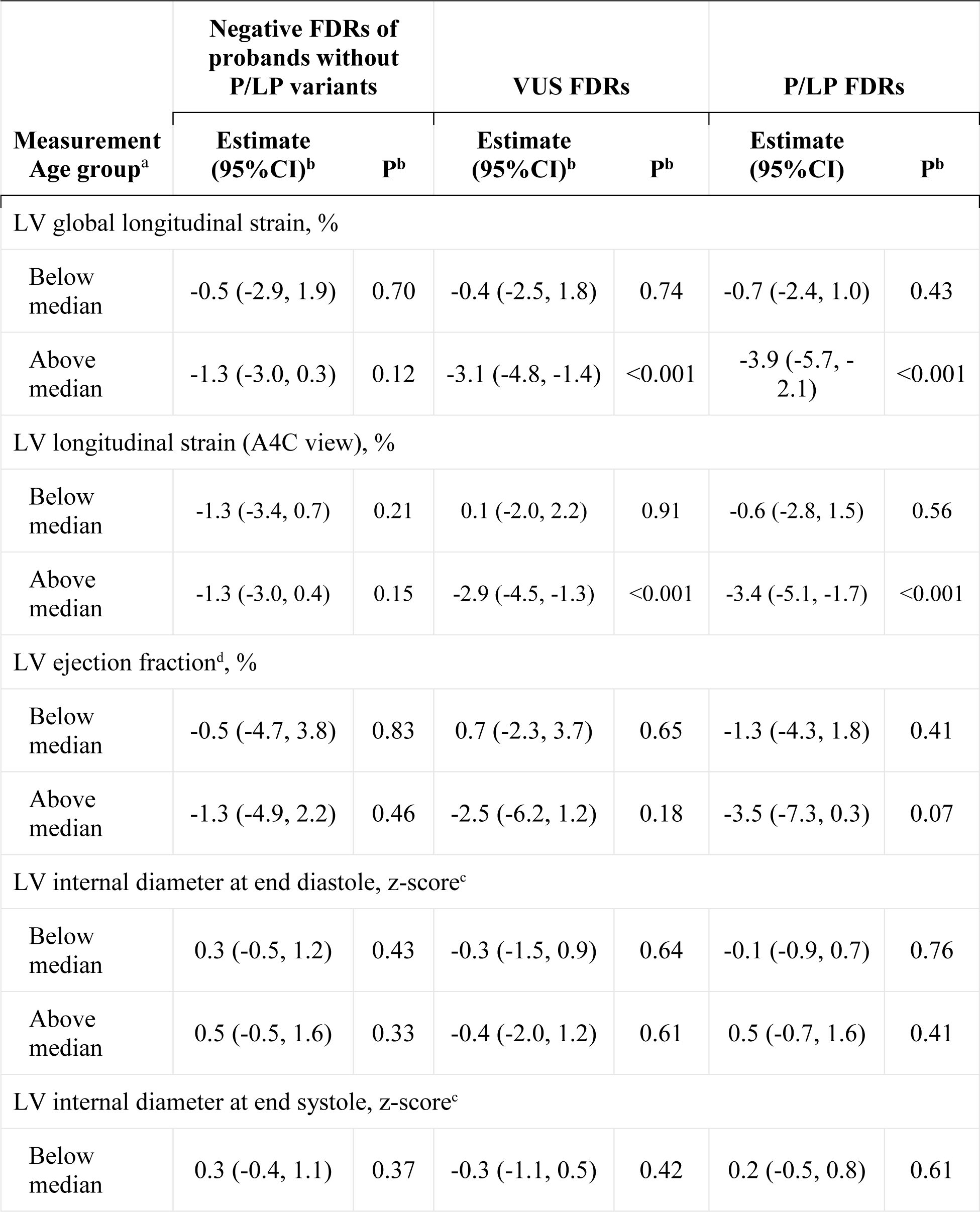
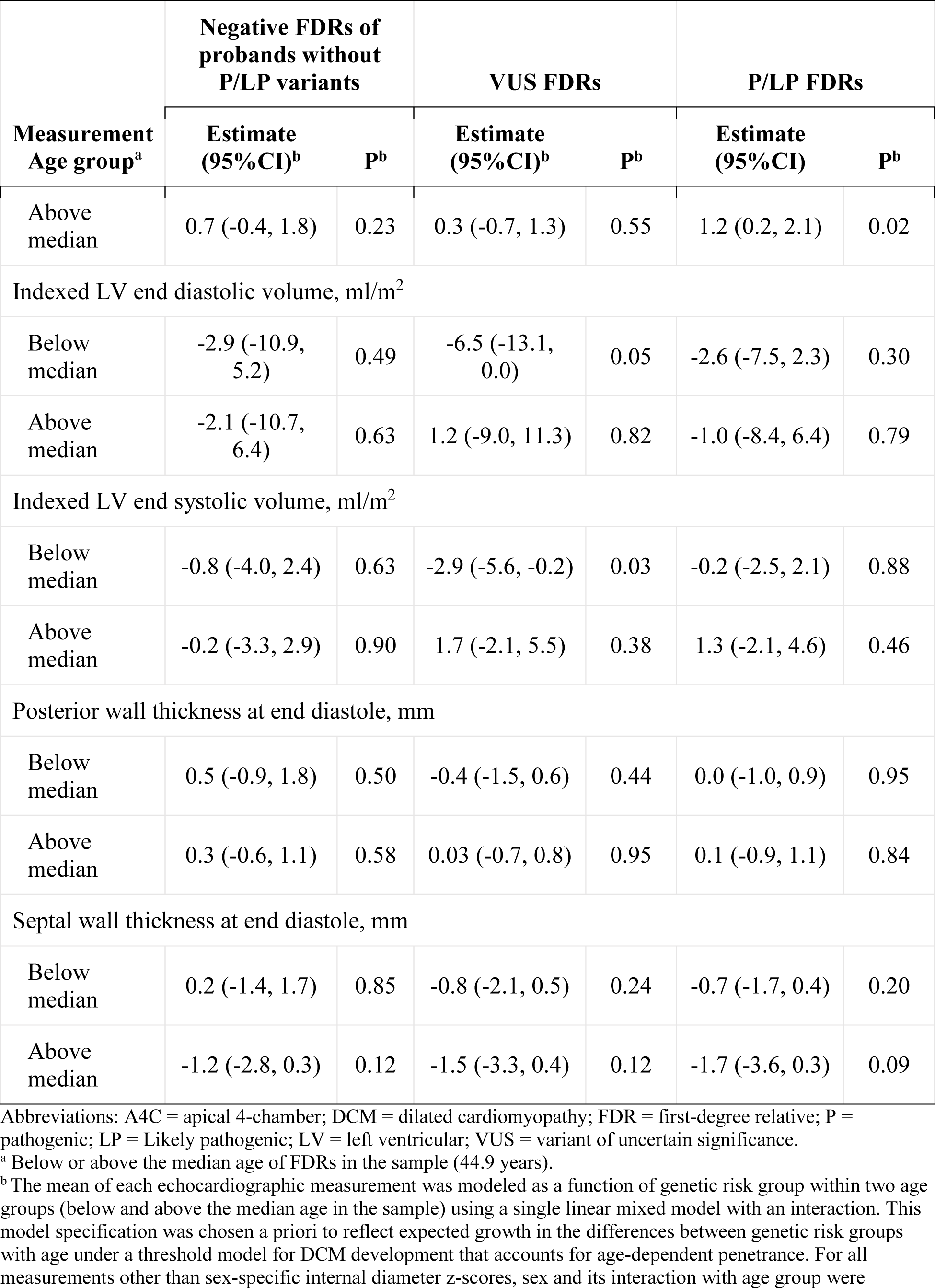
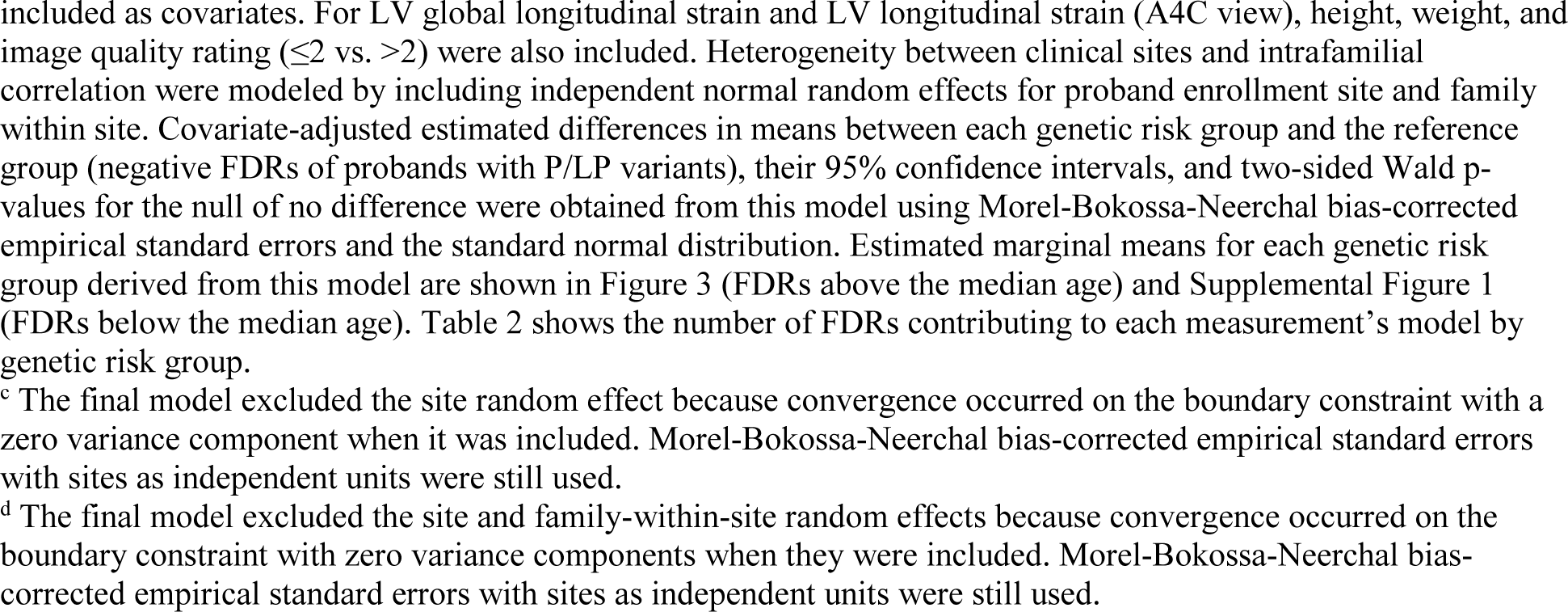
Covariate-adjusted estimated mean differences in echocardiographic measurements between other genetic risk groups and the reference group

| Measurement<br>Age group<sup>a</sup> | Negative FDRs of<br>probands without<br>P/LP variants |  | VUS FDRs |  | P/LP FDRs |  |
| --- | --- | --- | --- | --- | --- | --- |
|  | Estimate<br>(95%CI)<sup>b</sup> | P<sup>b</sup> | Estimate<br>(95%CI)<sup>b</sup> | P<sup>b</sup> | Estimate<br>(95%CI) | P<sup>b</sup> |
| LV global longitudinal strain, % |  |  |  |  |  |  |
| Below<br>median | -0.5 (-2.9, 1.9) | 0.70 | -0.4 (-2.5, 1.8) | 0.74 | -0.7 (-2.4, 1.0) | 0.43 |
| Above<br>median | -1.3 (-3.0, 0.3) | 0.12 | -3.1 (-4.8, -1.4) | <0.001 | -3.9 (-5.7, -2.1) | <0.001 |
| LV longitudinal strain (A4C view), % |  |  |  |  |  |  |
| Below<br>median | -1.3 (-3.4, 0.7) | 0.21 | 0.1 (-2.0, 2.2) | 0.91 | -0.6 (-2.8, 1.5) | 0.56 |
| Above<br>median | -1.3 (-3.0, 0.4) | 0.15 | -2.9 (-4.5, -1.3) | <0.001 | -3.4 (-5.1, -1.7) | <0.001 |
| LV ejection fraction<sup>d</sup>, % |  |  |  |  |  |  |
| Below<br>median | -0.5 (-4.7, 3.8) | 0.83 | 0.7 (-2.3, 3.7) | 0.65 | -1.3 (-4.3, 1.8) | 0.41 |
| Above<br>median | -1.3 (-4.9, 2.2) | 0.46 | -2.5 (-6.2, 1.2) | 0.18 | -3.5 (-7.3, 0.3) | 0.07 |
| LV internal diameter at end diastole, z-score<sup>c</sup> |  |  |  |  |  |  |
| Below<br>median | 0.3 (-0.5, 1.2) | 0.43 | -0.3 (-1.5, 0.9) | 0.64 | -0.1 (-0.9, 0.7) | 0.76 |
| Above<br>median | 0.5 (-0.5, 1.6) | 0.33 | -0.4 (-2.0, 1.2) | 0.61 | 0.5 (-0.7, 1.6) | 0.41 |
| LV internal diameter at end systole, z-score<sup>c</sup> |  |  |  |  |  |  |
| Below<br>median | 0.3 (-0.4, 1.1) | 0.37 | -0.3 (-1.1, 0.5) | 0.42 | 0.2 (-0.5, 0.8) | 0.61 |

| Measurement<br>Age group<sup>a</sup> | Negative FDRs of probands without P/LP variants |  | VUS FDRs |  | P/LP FDRs |  |
| --- | --- | --- | --- | --- | --- | --- |
|  | Estimate (95%CI)<sup>b</sup> | P<sup>b</sup> | Estimate (95%CI)<sup>b</sup> | P<sup>b</sup> | Estimate (95%CI) | P<sup>b</sup> |
| Above median | 0.7 (-0.4, 1.8) | 0.23 | 0.3 (-0.7, 1.3) | 0.55 | 1.2 (0.2, 2.1) | 0.02 |
| Indexed LV end diastolic volume, ml/m<sup>2</sup> |  |  |  |  |  |  |
| Below median | -2.9 (-10.9, 5.2) | 0.49 | -6.5 (-13.1, 0.0) | 0.05 | -2.6 (-7.5, 2.3) | 0.30 |
| Above median | -2.1 (-10.7, 6.4) | 0.63 | 1.2 (-9.0, 11.3) | 0.82 | -1.0 (-8.4, 6.4) | 0.79 |
| Indexed LV end systolic volume, ml/m<sup>2</sup> |  |  |  |  |  |  |
| Below median | -0.8 (-4.0, 2.4) | 0.63 | -2.9 (-5.6, -0.2) | 0.03 | -0.2 (-2.5, 2.1) | 0.88 |
| Above median | -0.2 (-3.3, 2.9) | 0.90 | 1.7 (-2.1, 5.5) | 0.38 | 1.3 (-2.1, 4.6) | 0.46 |
| Posterior wall thickness at end diastole, mm |  |  |  |  |  |  |
| Below median | 0.5 (-0.9, 1.8) | 0.50 | -0.4 (-1.5, 0.6) | 0.44 | 0.0 (-1.0, 0.9) | 0.95 |
| Above median | 0.3 (-0.6, 1.1) | 0.58 | 0.03 (-0.7, 0.8) | 0.95 | 0.1 (-0.9, 1.1) | 0.84 |
| Septal wall thickness at end diastole, mm |  |  |  |  |  |  |
| Below median | 0.2 (-1.4, 1.7) | 0.85 | -0.8 (-2.1, 0.5) | 0.24 | -0.7 (-1.7, 0.4) | 0.20 |
| Above median | -1.2 (-2.8, 0.3) | 0.12 | -1.5 (-3.3, 0.4) | 0.12 | -1.7 (-3.6, 0.3) | 0.09 |

Abbreviations: A4C = apical 4-chamber; DCM = dilated cardiomyopathy; FDR = first-degree relative; P = pathogenic; LP = Likely pathogenic; LV = left ventricular; VUS = variant of uncertain significance.
<sup>a</sup> Below or above the median age of FDRs in the sample (44.9 years).
<sup>b</sup> The mean of each echocardiographic measurement was modeled as a function of genetic risk group within two age groups (below and above the median age in the sample) using a single linear mixed model with an interaction. This model specification was chosen a priori to reflect expected growth in the differences between genetic risk groups with age under a threshold model for DCM development that accounts for age-dependent penetrance. For all measurements other than sex-specific internal diameter z-scores, sex and its interaction with age group were
<sup>c</sup> The final model excluded the site random effect because convergence occurred on the boundary constraint with a zero variance component when it was included. Morel-Bokossa-Neerchal bias-corrected empirical standard errors with sites as independent units were still used.
<sup>d</sup> The final model excluded the site and family-within-site random effects because convergence occurred on the boundary constraint with zero variance components when they were included. Morel-Bokossa-Neerchal bias-corrected empirical standard errors with sites as independent units were still used.

## DISCUSSION

In a multicenter study of unaffected FDRs of probands with DCM, FDRs above the sample median age who harbored either P/LP variants or VUSs had lower absolute values of LV GLS, an early measure of abnormal cardiac mechanics, relative to negative FDRs of probands both with and without P/LP variants. These differences were attenuated in FDRs below the median age in the sample, which is consistent with the age-dependent nature of DCM development. Moreover, the observation that older FDRs with only VUSs had lower absolute LV GLS values than both the reference group and another group of FDRs with uncertain genetic risk provides the first evidence that some VUSs are clinically relevant for DCM. Taken together, these findings suggest that a reduction in absolute LV GLS values may provide an opportunity for earlier detection of incipient DCM among FDRs who are genetically at risk but still clinically unaffected (Graphical Abstract), supporting a long-term goal of the DCM Research Project to identify and rigorously define a pre-DCM phenotype in at-risk FDRs with longitudinal observation.

GLS has been well established for its prognostic value in multiple cardiovascular disease states including ischemic heart disease^23^ and all cause HF.^24^ For DCM, GLS has mostly been studied for improved risk prediction in patients with an existing DCM phenotype (e.g., LVEF <50% or imaging evidence of myocardial fibrosis).^25^ Our work builds on prior single-center studies that also evaluated the role of GLS to detect subclinical LV dysfunction in family members of DCM probands.^12–14^ One study demonstrated reduced strain parameters in genotype-positive phenotype-negative family members when compared with controls.^13^ Another demonstrated reduced LV GLS among variant-positive family members, but did not evaluate VUSs, and the convenience sample was on average a decade older than our study with co-morbid conditions.^12^

While differences in LV GLS were seen in this study between genetic risk groups, overall absolute mean values were still within the normal range, and only a small fraction of the study population had borderline values of 16-18% (n=14, 12.1%) or abnormal values of <16% (n=3, 2.6%). Nonetheless, the results highlight observable differences among clinically normal at-risk FDRs that support the hypothesis that a pre-DCM phenotype based on LV GLS emerges with advancing age in a manner dependent upon genetic risk. A larger longitudinal study is needed to offer greater insight into these observed phenomena.

The finding that older FDRs harboring only their proband’s VUSs also showed a reduction in LV GLS is an important and novel observation. Per usual genetic cardiomyopathy guidelines,^3,5^ VUSs are not recommended to be used for clinical decision making and are strongly discouraged for use for predictive testing for family risk stratification. This is because an at-risk individual could be released from ongoing clinical surveillance while still harboring other unknown variants that incur genetic risk. Such caution is appropriate, and the authors agree with this approach. Variant classification categories have been developed within a single gene, Mendelian disease framework,^4^ where one major variant of strong effect (P/LP) segregates with a well-recognized phenotype such as DCM throughout a kindred, as previously reviewed in detail.^9^ Thus, P/LP classifications require a high level of confidence that the variant causes nearly all the disease in the kindred; if such confidence is not warranted by available evidence, the variant is classified as a VUS.

Issues and approaches to VUS resolution are well established, with recognition that clarifying clinical relevance, if any, of many VUSs may take years.^26^ Considerable effort has been expended to resolve possible clinical effects of VUSs,^27,28^ with a finding that 89% of missense variants classified as a VUS for Lynch syndrome were suggested to be functionally neutral.^28^ In contrast, a study of pediatric DCM patients that included a retrospective reclassification of VUSs found that only 29% were reclassified as likely benign variants.^29^ Regardless, as shown here and previously in a smaller data set,^30^ VUSs rigorously and conservatively classified^6^ by our studies were associated with worse LV structure and function among relatives.

While from this study one might infer intermediate pathogenicity for most DCM VUSs, it is likely that VUSs range in impact from little to no effect to fully pathogenic, with the latter lacking case or other data to be elevated from VUS to LP or P.^6,9^ Also, more than one P/LP/VUS variant has been observed in 21% of DCM probands,^6^ suggesting that the Mendelian/single gene model upon which current variant adjudication standards are based^4^ may be incomplete for DCM.^6,9,30^ As noted above and previously discussed,^6,9,30^ deciphering DCM genetic complexity, including clarification of the DCM disease model and role of variants now classified as VUSs, is necessary for a more complete approach to DCM clinical genetics care.

### Limitations

This report has limitations. First, in this sample, results could only be obtained for FDRs of probands with predominantly European ancestry; a planned larger analysis of the more diverse full cohort^8^ will resolve this important issue. Nevertheless, this study provided novel insight into how genetic risk manifests with age among an initial sample of clinically unaffected FDRs of DCM probands. Moreover, while this study has shown that VUSs were associated with worse LV GLS in clinically unaffected FDRs, the association does not necessarily reflect the direct causal effect of harboring a VUS on LV GLS. Even so, the observed association still establishes harboring a VUS as a clinically relevant indicator of elevated DCM risk. Future work in the larger data set from the full cohort may also provide more specificity regarding the impact of VUSs by gene or variant ontology, which will provide more helpful clinical information. Finally, the associations presented in this study are cross-sectional; longitudinal measurements will be required to describe and compare age trajectories of cardiac mechanics in FDRs with varying levels of genetic risk.

## Conclusion

This multi-center study provided evidence suggesting that LV cardiac mechanics, as measured by GLS, worsen more rapidly with age in FDRs without LVSD or LVE who have higher levels of genetic risk. Moreover, worse LV GLS was observed not only among older FDRs with P/LP variants but also among those with VUSs, suggesting that some DCM-relevant VUSs are clinically relevant. These findings suggest that declines in LV GLS could define a pre-DCM phenotype that may provide opportunity for identification of at-risk individuals earlier in the DCM disease process.

## Data Availability

Data used to produce the findings in the present study are available from the corresponding author upon reasonable request.

## ACKNOWLEDGEMENTS

The investigators thank the families with DCM who have participated in this study, without whom this effort would not be possible. The DCM Precision Medicine Study was supported by computational infrastructure provided by The Ohio State University Division of Human Genetics Data Management Platform and the Ohio Supercomputer Center.

