## Supporting Information for "Differences in Cardiac Mechanics among Genetically At-Risk First-Degree Relatives: The DCM Precision Medicine Study"

### Supplemental Methods

#### Ancestry Inference

Genotyping

Array-based genotyping was performed at the University of Washington Northwest Genomics Center (NWGC) for all DCM Research Project samples that underwent exome sequencing during the study period, including some from studies other than the DCM Precision Medicine Study. Samples were normalized using a PerkinElmer Janus Workstation in preparation for the Illumina Infininium HD genotyping assay using the Global Screening Array (GSA) v1.0 and v2.0 beadchip. Completed assays were scanned on an Illumina iScan, and image files were transferred to the NWGC network to perform analysis. FinalReport files for genotyping batches were generated using Illumina GenomeStudio software (v2.0.3) with the appropriate manifest files (GSA-24v1-0_C1 or GSA-24v2-0_A1). Samples with a call rate <97% were considered failed and excluded from the batch FinalReport file.

Initial Quality Control

Illumina product and support files, including the manifest, were downloaded from the Illumina support site (https://support.illumina.com/array/array_kits/infinium-global-screening-array/downloads.html). These files were used to identify 607,189 biallelic single-nucleotide polymorphisms (SNPs) that appeared on both GSA versions and were not coordinate duplicates. Batch Illumina FinalReports for each array version were combined into a PLINK LGEN fileset, and PLINK (version 1.90b6.21)[1] was used to create binary filesets, flipping allele strand as necessary so that all alleles were expressed on the same strand as the GRCh37 reference sequence. These filesets were then merged into a single binary fileset using PLINK.

Initial SNP and sample quality control was completed using PLINK. A total of 19,739 SNPs with call rates <98%[2] among eligible subjects or with call rates differing by >2% between array versions were removed. No subjects with genotype call rates <97% among remaining SNPs had to be removed. One SNP having Mendel errors in more than 5% of nuclear families was excluded; no nuclear families had to be removed due to Mendel errors in more than 1% of SNPs. Genotypes for any remaining Mendel errors were set to missing.

Principal Components Analysis

Subsequent quality control steps followed published recommendations to use ancestry-adjusted quality control metrics in diverse cohorts.[3] The 1000 Genomes Phase 3 integrated callset[4] available at http://ftp.1000genomes.ebi.ac.uk/vol1/ftp/release/20130502/ was used to obtain ancestry principal components onto which DCM Research Project samples could be projected for subsequent steps. All analyses using R or Bioconductor packages were performed in R version 4.1.1 (R Foundation).

*Joint Fileset Assembly*

High-quality, common autosomal SNPs passing previous QC steps with call rates of at least 99.5% in probands and minor allele frequencies (MAFs) ≥5% in probands were selected from the DCM Research Project cohort. For each 1000 Genomes Phase 3 autosomal VCF, bcftools (version 1.15.1) was used to drop sample NA20318 due to relatedness and select biallelic SNPs with:

1) CHROM, POS, REF, and ALT matching a GSA SNP selected from the DCM Research project cohort;
2) FILTER=. or FILTER=PASS;
3) Call rate ≥99.5%; and
4) MAF ≥5%

The resulting compressed BCFs were then concatenated with bcftools into a single compressed BCF. The compressed BCF was then converted to a PLINK binary fileset and merged with the GSA ancestry SNP fileset using PLINK to create a joint fileset with 4,033 individuals and 267,947 SNPs.

*Identification of Unrelated Subjects*

The KING-robust algorithm[5] implemented in the SNPRelate Bioconductor package[6] (version 1.28.0) was used to estimate kinship coefficients between pairs of individuals in the joint fileset. KING-robust does not assume linkage equilibrium between markers and is robust to departures from Hardy-Weinberg equilibrium (HWE) in the direction of excess heterozygosity;[5] the KING manual specifically recommends against performing any type of linkage disequilibrium (LD) pruning.

Only 1000 Genomes samples that were unrelated to every DCM sample in this joint fileset (kinship coefficient <0.025)[7] were used for ancestry analyses. This served to ensure that none of the DCM Research Project samples were related to the 1000 Genomes samples used as a reference so that the online augmentation, decomposition, and Procrustes approach could be used for projection of our study samples onto the 1000 Genomes principal components.[8,9] Among the remaining 1000 Genomes samples, the PC-AiR algorithm[7] implemented in the pcairPartition function of the GENESIS Bioconductor package (version 2.24.2) was used to select an unrelated subset with the most ancestral divergence. We used the kinship cutoff given above and a divergence cutoff of -0.025 as recommended.[7] We used the same PC-AiR approach to identify an unrelated subset of DCM Research Project probands for a subsequent structured HWE analysis. After filtering, there were 2,422 subjects from 1000 Genomes unrelated to each other and any of the eligible DCM Research Project subjects who could be used for structured HWE and ancestry analysis and 1,266 unrelated DCM Research Project probands who could be used for structured HWE analysis. Note that these probands include individuals enrolled in other DCM Research Project studies who underwent exome sequencing.

*Structured Hardy-Weinberg Equilibrium Testing*

The structured HWE method of Hao and Storey (2019)[10] implemented the lfa Bioconductor package tests for deviation from HWE after removing the effects of population structure and was used to test SNPs that could potentially be used in ancestry analysis for deviation from HWE. A recent version on GitHub (2.0.8; d2ced17), rather than the one on Bioconductor, was used to enable BEDMatrix (version 2.0.3) support. These tests were performed separately in unrelated probands and unrelated 1000 Genomes reference samples to identify poor-quality SNPs for each genotyping technology (GSA or 1000 Genomes integrated calling). The sHWE function from lfa was not used directly but was reimplemented based on the source code in a way that improved memory efficiency and parallelism.

The number of logistic factors, $k$, was selected adaptively by the p-value entropy approach in Algorithm 2 of Hao and Storey (2019) with 150 bins.[10] In particular, $k$ was incremented and the p-value entropy among all but the smallest bin calculated. Once the entropy remained at or below the value with $k$ logistic factors for $k + 1$ and $k + 2$ logistic factors, analysis proceeded with $k$ logistic factors. P-values were calculated by simulating data for each SNP under the null model given the estimated individual-specific allele frequencies and calculating the test statistic, which effectively gave a bootstrap sample of size equal to the number of markers under the null hypothesis because the test statistic is a pivotal quantity.[10] Empirical p-values were calculated using the formula that counts the observed test statistic in both the numerator and denominator.[11]

The structured HWE p-values obtained with this $k$ were used to estimate SNP q-values as well as the proportion of SNPs in HWE, $\pi_0$, using the bootstrap method implemented in the Bioconductor qvalue package (version 2.26.0).[10,12] Strong evidence of deviation from HWE was defined as having a q-value ≤0.05 because no more than 5% of SNPs meeting this threshold would be expected to actually be in HWE, including in the presence of local dependence induced by LD.[12] While no SNPs met the q-value threshold in unrelated DCM Research Project probands, a total of 4,261 SNPs with relatively strong evidence of deviation from HWE in unrelated 1000 Genomes reference samples were eliminated for subsequent ancestry analyses, leaving 263,686 SNPs.

*Principal Components Analysis of 1000 Genomes Reference Samples*

Principal components analysis (PCA) was performed on the 2,422 1000 Genomes reference samples found to be unrelated to each other and all DCM Research Project samples. The bigsnpr R package (version 1.10.8) and its dependencies were used to perform the analysis.[9] SNPs located in long-range LD regions identified in Table S13 of Bycroft et al. (2018)[13] were eliminated from those in sHWE, and the remainder were subjected to a round of LD clumping using the bigsnpr bed_clumping function with default parameters ($r^2 > 0.2$ within a 500 kb window) to remove those in LD, leaving 130,676 SNPs for use in principal components analysis. Because the reference set contained 26 populations, including some that were admixed, these should be represented by at most 25 large eigenvalues based on population genetics arguments,[14] and 26 principal components (PCs) was chosen as a safe upper bound for the truncated singular value decomposition (SVD).

Outlier SNPs were detected based the loading vector's robust Mahalanobis distance from the centroid. While Privé et al. (2020)[9] used the tukey_mc_up function in the bigutilsr R package to determine this by adjusting the coefficient for multiplicity, the parameters for the medcouple-adjusted boxplot fence were derived for a coefficient of 1.5 only,[15] so it is unclear whether this approach would achieve the desired type I error. Based on Hubert and Vandervieren (2008), approximately 0.35% of SNPs should fall outside of the upper fence of the medcouple-adjusted boxplot if they are all independent draws from the same distribution. Because these are approximations, outlier SNPs were defined as those with distances larger than the 0.5% of SNPs outside of the upper fence with the smallest distances. If less than 0.5% of SNPs were outside of the upper fence, there were no outliers. For example, if there were 100,000 SNPs and 531 outside of the outer fence, the 31 of the 531 with the largest robust Mahalanobis distances would be considered outliers. If there were only 479 SNPs outside of the upper fence, there would be no outliers.

Outlier samples in the PC score space were identified using Probabilistic Local Outlier Factor approach.[9] We used PC scores scaled to have the same variance ($U$ from the SVD)[16] rather than the PC scores themselves ($UD$ from the SVD), although the latter are the coordinates actually used when plotting individuals in PC space. This scaling ensures that each PC has equal weight in sample outlier calculations rather than giving large weights to the first few PCs. Cutpoints for the statistic were chosen based upon visual inspection of the histogram and pairwise PC score plots in each iteration.

PCA was performed iteratively according to the steps below until no further outlier SNPs or samples were identified:

1) Perform SVD subiterations:
    a. Perform SVD using the bigsnpr bed_randomSVD function with the current list of included samples and SNPs.

b. Calculate the robust Mahalanobis distance of the loading vector across PCs for each SNP using the dist_ogk function in the bigutilsr R package (version 0.3.4).[9]
c. Identify and remove outlier SNPs. If none are found, stop subiterations.

2) Identify and remove outlier reference samples.

This iterative process led to the removal of 153 SNPs and 16 reference samples, leaving 130,523 SNPs and 2,406 reference samples for the final PCA. On the basis of the scree plot and pairwise PC plots from the final PCA, the first 20 PCs were adequate to capture all relevant variation between populations in the 1000 Genomes reference sample. This conclusion squares well with the $k = 21$ logistic factors arrived at by a different method in the logistic factor analysis for structured HWE above. SNP loading plots for each PC showed a similar distribution across the genome without any localized regions of high loadings, suggesting that all PCs were capturing population structure.

*Projection of DCM Research Project Samples onto 1000 Genomes Principal Components*

The Online Augmentation, Decomposition, and Procrustes (OADP) approach[8] as implemented in the bed_projectSelfPCA function of the bigsnpr package was used to obtain predicted PC scores for DCM Research Project samples on the 1000 Genomes PCs. Advantages of using projected scores based on OADP include:

1) Predicted PC scores for an individual are the same regardless of who is included in the sample for prediction, which obviates the need to perform a second PCA after sample quality control and allows for easy addition of additional subjects without changing prior results.

2) Projection can be used to obtain PC scores for related individuals.[7] With OADP, relatedness in the study samples should not impact prediction accuracy as long as reference samples are unrelated to each other[8] and the study samples.[9]
3) OADP eliminates centroid bias in predicted PC scores that can occur with standard techniques.[8,9]

Ancestry-Adjusted Quality Control

On the basis of the 1000 Genomes PCA, adjustment for 20 PCs in DCM Research Project samples, all of which should be from populations included in the 1000 Genomes reference sample or mixtures of them, was deemed sufficient to remove the effects of population structure for ancestry-adjusted quality control measures.

*Autosomal Heterozygosity*

In order to identify and remove potentially poor quality samples in terms of heterozygosity, an approach to account for ancestry similar to the one detailed in Bycroft et al. (2018) was employed.[13] Raw autosomal heterozygosity for markers used in the PCA was estimated in the 1000 Genomes reference set, and an ordinary least squares regression of raw heterozygosity on an intercept and a bias term representing the effect of population structure was fit. The bias term included linear, quadratic, and two-way interacted PC score variables for the first 20 PC scores and had the same functional form as the bias term used in Bycroft et al. (2018).

Raw autosomal heterozygosity and its expected value based on the regression model fit to the 1000 Genomes reference samples were calculated in the projected DCM Research Project samples. Samples with observed heterozygosity more than 3 SDs (in terms of model root mean squared error) above their expected value based on ancestry were considered to have unusually high heterozygosity. In order to confirm that samples with unusually low heterozygosity were not subject to poor quality genotyping, a plot of the relationship of the difference between observed and expected heterozygosity and the total length of all long runs of homozygosity was also produced. The total length of all long runs of autosomal homozygosity (segments greater than or equal to 1000 kb) for each projected DCM sample with the same SNP set was calculated using PLINK with the --homozyg-kb 1000 option and all other settings left as their default values (i.e., --homozyg-density 50, --homozyg-gap 1000, --homozyg-window-snp 50, --homozyg-window-het 1, --homozyg-window-missing 5, --homozyg-window-threshold 0.05 and no limit on the number of heterozygous calls per run of homozygosity).

A total of 8 projected DCM Research Project samples were considered outliers due to high autosomal heterozygosity. Six samples had evidence of recent ancestral admixture that would explain the high heterozygosity, but the remaining 2 were removed from further processing. The expected negative relationship between PC-corrected heterozygosity and the total length of all runs of homozygosity was confirmed; thus, no samples were excluded for having unusually low heterozygosity.

*Sex Discordance*

To check for potential sample swaps based on discordance from reported sex, X chromosome heterozygosity and the Y chromosome genotype missing rate were computed for the remaining projected samples. Raw X chromosome heterozygosity was plotted against the Y chromosome genotype missing rate with samples faceted by self-reported biological sex, and the plot was visually inspected to identify any potential sex-related discordance. There appeared to be no sex discordance among the samples considered.

*Expected Relatedness*

As the DCM Research Project studies are family-based, many families had genotype data on multiple members included in the sample set. To verify that the pedigree-based relatedness between samples was correct, identify potential sample swaps, and identify cryptically related probands, pairwise kinship coefficients and pairwise probabilities of sharing zero alleles identical by descent ($k_0$) were obtained using the PC-Relate algorithm implemented in the pcrelate function of the GENESIS Bioconductor package with the same set of SNPs used in PCA.[17] PC-Relate accounts for diverse ancestry among samples by modeling the individual-specific allele frequency at a SNP as a function of ancestry representative PCs and then using the estimated individual-specific allele frequencies from this model in lieu of the sample allele frequencies in modified kinship and $k_0$ estimators [17]. The resulting estimates should therefore reflect relatedness due only to recent pedigree structure.[17] The combined set containing the 2,406 1000 Genomes reference samples used for PCA was used as a training set, and pairwise relatedness estimates were calculated for all DCM Research Project samples that passed all previous quality control steps. Individual-specific allele frequency estimation was based on 20 PCs, and, if an estimated individual-specific minor allele frequency at a SNP was less than 0.01, that SNP was excluded from the analysis for that individual. Overall genotype scaling was used in the kinship estimator.

Once the estimated kinship coefficients and $k_0$ values were calculated, pairwise estimated kinship coefficients were plotted against the $k_0$ values for each type of pedigree-based relationship. The inference criteria of Manichaikul et al. (2010)[5] for kinship and $k_0$ were used where possible to determine whether estimated relatedness matched the expectation based on the pedigree relationship. For relationships not falling under these inference criteria, QuickPed (https://magnusdv.shinyapps.io/quickped/)[18] was used to calculate expected kinship and $k_0$. Upon visual inspection, most pairwise combinations of estimated kinship and $k_0$ met the inference criteria or expectation for the reported pedigree relationship, although 9 pairwise combinations did not. Each of these cases was investigated, and it was determined that 4 DCM samples were likely involved in accidental sample swaps. The potentially swapped samples were removed from subsequent analyses. In addition, three probands who were 3rd degree relatives or closer to previously enrolled probands were identified.

Ancestry Estimation for 1000 Genomes Reference Samples

ADMIXTURE (version 1.3.0) was used to estimate global ancestry proportions and allele frequencies in the inferred ancestral populations for the 2,406 unrelated 1000 Genomes reference samples used in PCA.[19-21] Note that the ADMIXTURE likelihood, like PCA, assumes unrelated reference samples,[19] and inclusion of related samples can lead to lower-quality ancestral allele frequency estimates.[21] As ADMIXTURE also assumes linkage equilibrium between markers, the LD-pruned SNP set from the final PCA was used, as recommended.[19] ADMIXTURE relies on the calling user to provide a value, $K$, which represents the number of ancestral populations believed to be among the input samples.

For a given $K$, the ADMIXTURE likelihood has $K!$ equivalent global maxima in which the ancestry proportions and allele frequencies are identical but appear in different columns of $Q$ and rows of $P$.[19] As ADMIXTURE uses a greedy, hill-climbing search to maximize the likelihood, 30 runs from different starting values were performed for each $K$ by varying the random seed[22] to have a greater chance of achieving a well-supported global maximum. The maximal runs with a maximized loglikelihood numerically equivalent to the maximum achieved across all runs (i.e., absolute value of relative difference <3e-8) for a given $K$ are likely to represent these equivalent global maxima as opposed to a local maximum attained due to poor initial values. To verify that the maximal runs represented the same solution up to a permutation of the columns of $Q$, pong (version 1.5)[23] was used to find the maximal weight alignments of columns in $Q$ for these runs and calculate their similarity for a given $K$. If the $Q$ matrices for the maximal runs in a given $K$ clustered into a single mode with high similarity, then the maximal runs were equivalent, and the putative global maximum was well-identified.

To determine the optimal $K$, ADMIXTURE's v-fold cross-validation procedure[20] with $v = 5$ was used to estimate the model's prediction error (deviance) for individual allele counts in each run. On the basis of these metrics (Supplemental Figure 2), $K = 9$ was selected because it had the lowest mean cross-validation error in a set of 13 equivalent maximal runs. Supplemental Figure 3 presents the estimated ancestry proportions for the maximal run with the highest loglikelihood for $K = 9$ with descriptive names for the inferred ancestral populations.

Projection of DCM Research Project Samples onto the 1000 Genomes Population Structure

To estimate the global ancestry proportions of the DCM Research Project samples, we used ADMIXTURE to project the DCM Research Project samples onto the 1000 Genomes population structure (i.e., allele frequencies) from the maximal run with the highest loglikelihood for $K = 9$.[21] Once again, 30 runs from different starting values were performed, and pong was used to verify that all maximal runs represented the same solution up to a permutation of the columns of $Q$. Ancestry proportion estimates for the DCM Research Project samples were obtained from the maximal run with the highest loglikelihood.

### Variant Analysis and Interpretation

Research exome sequencing was conducted and data processed as described previously[24] with the following modifications. The pipeline used by the NWGC was updated to use newer versions of key software components (BWA 0.7.10 to 0.7.15, Picard 1.111 to 2.6.0, and GATK 3.2-2/3.4-46 to 3.7-0) for the final batch of samples. Also, while joint calling at NWGC was done on smaller batches of samples during the study to allow continuous return of results, at study completion variants were jointly recalled across all DCM Research Project samples that underwent exome sequencing during the study period. Finally, the gnomAD[25] version in the annotation database in the Division of Human Genetics Data Management Platform was updated from release-170228 to 2.1.1 for the variant classifications used in the final analysis.

Rare protein-altering variants in 35 DCM genes underwent automated filtering followed by manual adjudication as described previously[24] with modifications described below. Variants were adjudicated using American College of Medical Genetics/Association of Molecular Pathologists (ACMG/AMP)[26] and the Clinical Genome Resource (ClinGen)-based criteria[27] tailored to DCM.[24] As part of this process, a computational algorithm first attempted to classify variant as benign, likely benign, unlikely to impact protein function, or low quality.[24] While the original algorithm used both QDFilter and SNPCluster VCF FILTER flags to classify variants as low quality, the SNPCluster filter was later eliminated due to false positives.

Variants that could not be computationally adjudicated were manually reviewed and certified for a final classification and confirmed with Sanger sequencing.[24] Modifications to the original criteria were approved by the Variant Adjudication Oversight Committee (VAOC) of The DCM Precision Medicine Study on August 4, 2022 and were applied to obtain the variant classifications used in the final analysis. These modifications included 1) elevating predicted loss of function (pLOF) variants located in the *DSP* gene from a moderate level of pathogenicity (PVS1_Moderate) to a strong level (PVS1_Strong) and 2) expanding the use of a strong level of evidence (PVS1_Strong) for pLOF variants in *TTN* beyond only the A-band to pLOF *TTN* variants in all highly expressed exons in the heart.

Both *DSP* and *TTN* are considered definitive evidence DCM genes by ClinGen.[28] *DSP* has a high probability of being LOF intolerant (gnomAD pLI = 1.00), and pLOF variants in *DSP* have been shown to be pathogenic and repeatedly demonstrated enrichment in DCM cases versus controls.[29-31] This enrichment that has been recapitulated in larger cohorts since initial preparation of the study variant adjudication criteria.[32] For *TTN* pLOF variants, over-representation in the A-band in DCM is also well established. Predicted LOF variants located in the distal I-band have also been shown to be enriched in cases versus controls.[33] A gene-first analysis further evaluated highly spliced in (percent spliced in [PSI] >0.9) *TTN* exons in the heart and demonstrated an increased odds of DCM with pLOF variants in high PSI *TTN* exons.[34] Therefore, the VAOC approved modifications to the study criteria to include expanding the use of PVS1 at a strong level of evidence of pLOF variants in high PSI exons of *TTN* beyond the A-band, including the Z, I, and exon 358 of the M-Band. To adopt a conservative, clinical classification approach, PVS1 was applied as strong evidence in pLOF variants in high PSI exons of *TTN* beyond the A-band only when the variant had been observed in at least one other unrelated DCM case and when no other P or LP variant(s) in other DCM-associated genes were present.

As some of these changes were fully implemented after generation of the sampling frame, a total of 3 first-degree relatives (FDRs) who harbored only variants of uncertain significance (VUSs) in the sampling frame were reclassified as harboring pathogenic or likely pathogenic (P/LP) variants for the final analysis.

### Supplemental Figures

**Supplemental Figure 1.** Echocardiographic measurements by genetic risk group in first-degree relatives below the median age in the sample

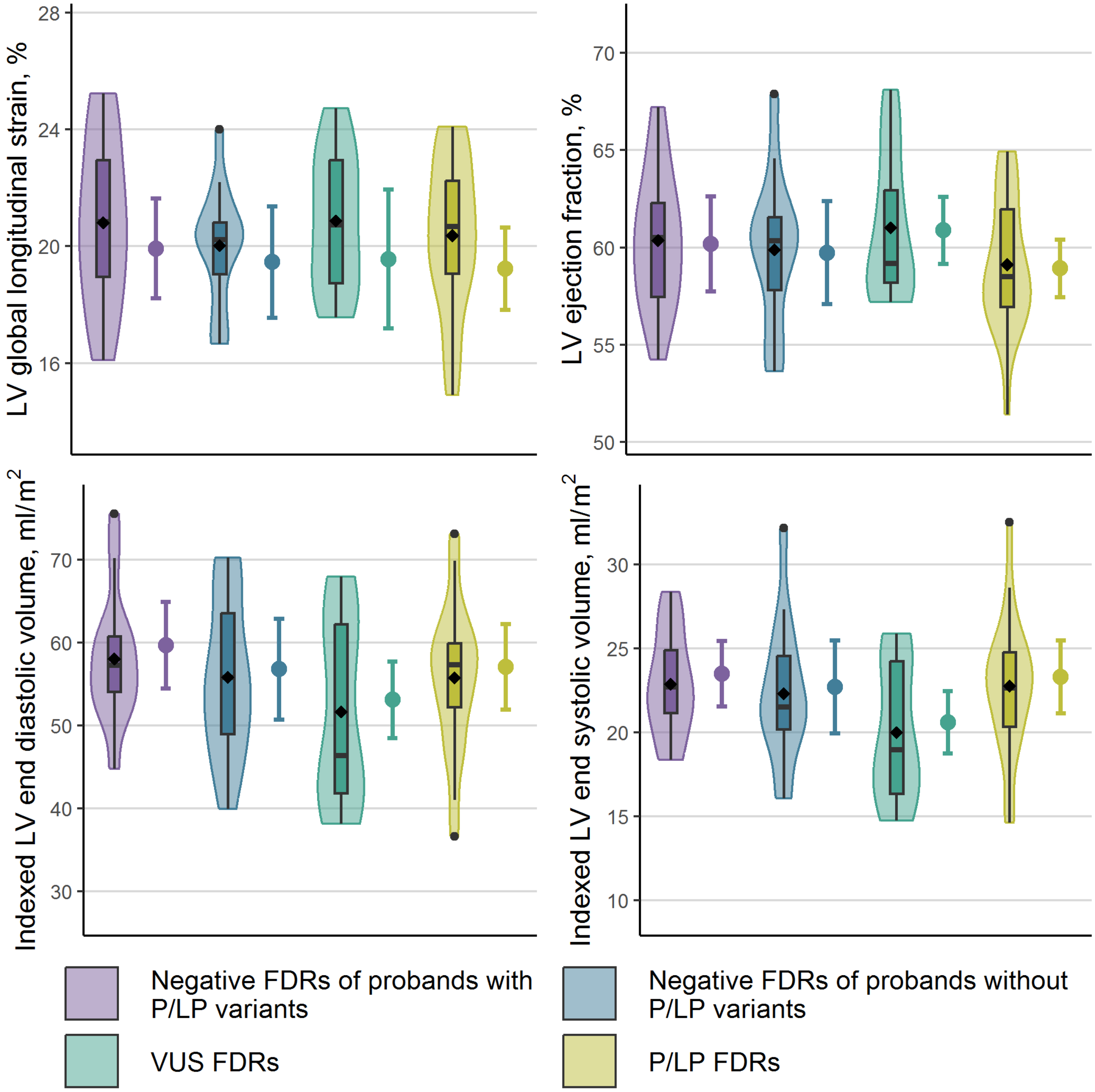

A violin plot with a superimposed box-and-whisker plot shows the distribution of the measurements among first-degree relatives (FDRs) in each genetic risk group, with a black diamond at the mean. Next to this, an interval plot shows the estimated marginal mean from the

linear mixed model analysis (point) as well as its 95% confidence interval (interval) obtained using Morel-Bokossa-Neerchal bias-corrected empirical standard errors and the standard normal distribution. For each genetic risk group, the estimated marginal mean is a covariate-adjusted estimate of the mean in a population of FDRs below the median age in the sample (44.9 years) that is half female. For LV global longitudinal strain, these populations also have the same mean height and weight and half image quality >2. Table 3 in the main text presents covariate-adjusted estimated mean differences between each genetic risk group and the reference group for FDRs below the median age from the same model; these are identical to the differences between the estimated marginal means in this figure. LP = likely pathogenic; LV = left ventricular; P = pathogenic; VUS = variant of uncertain significance.

**Supplemental Figure 2.** Maximized loglikelihoods and cross-validation error estimates for ADMIXTURE runs on 1000 Genomes reference samples for various *K*

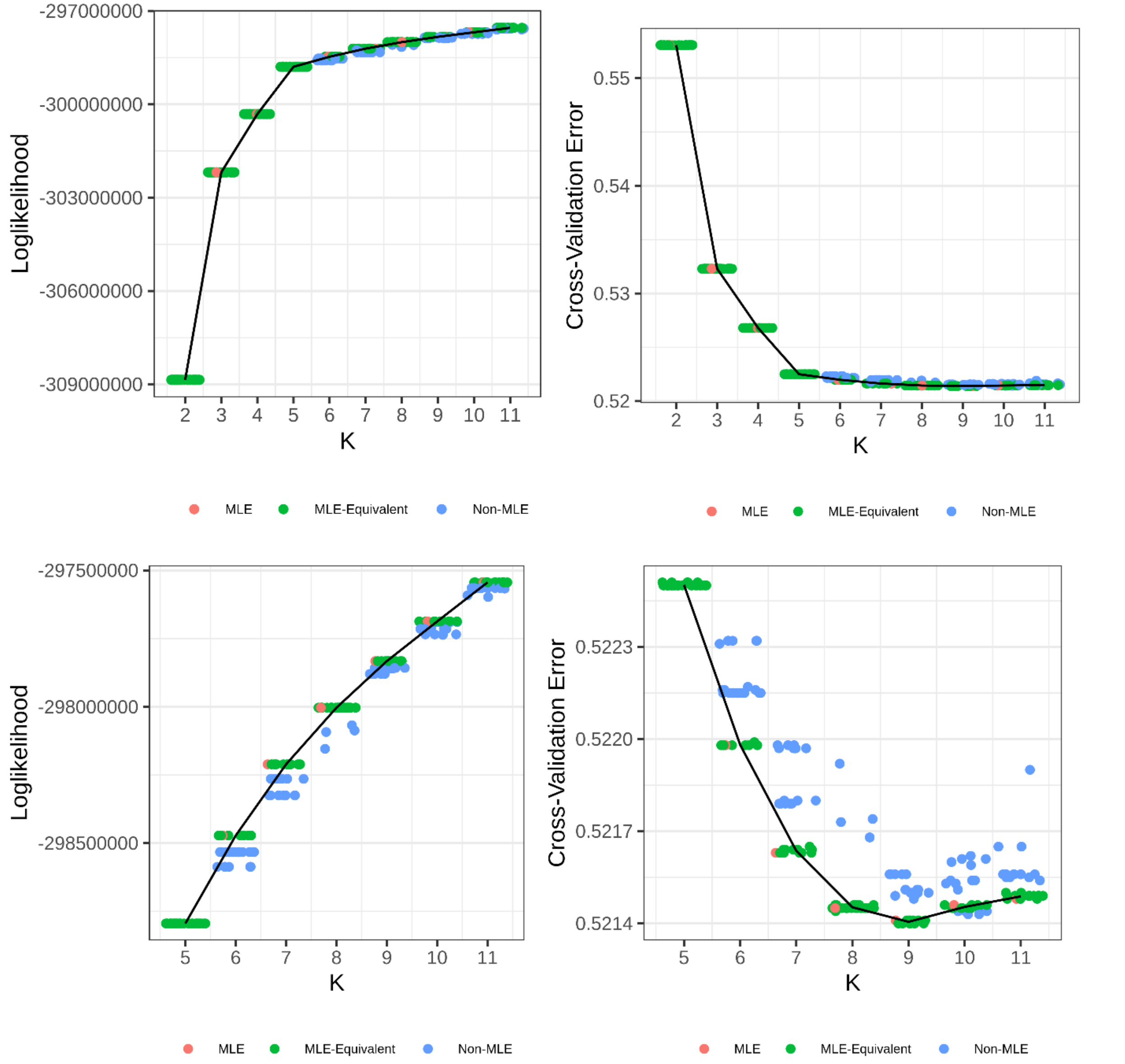

Each dot represents a single run; 30 independent runs with randomly chosen starting values were performed for each assumed number of ancestral populations (*K*). The black lines represent the mean of the vertical axis variable across the maximal runs (MLE and MLE-equivalent).

**Supplemental Figure 3.** Estimated global ancestry proportions for 1000 Genomes reference samples from the maximal run for the optimal number of assumed ancestral populations

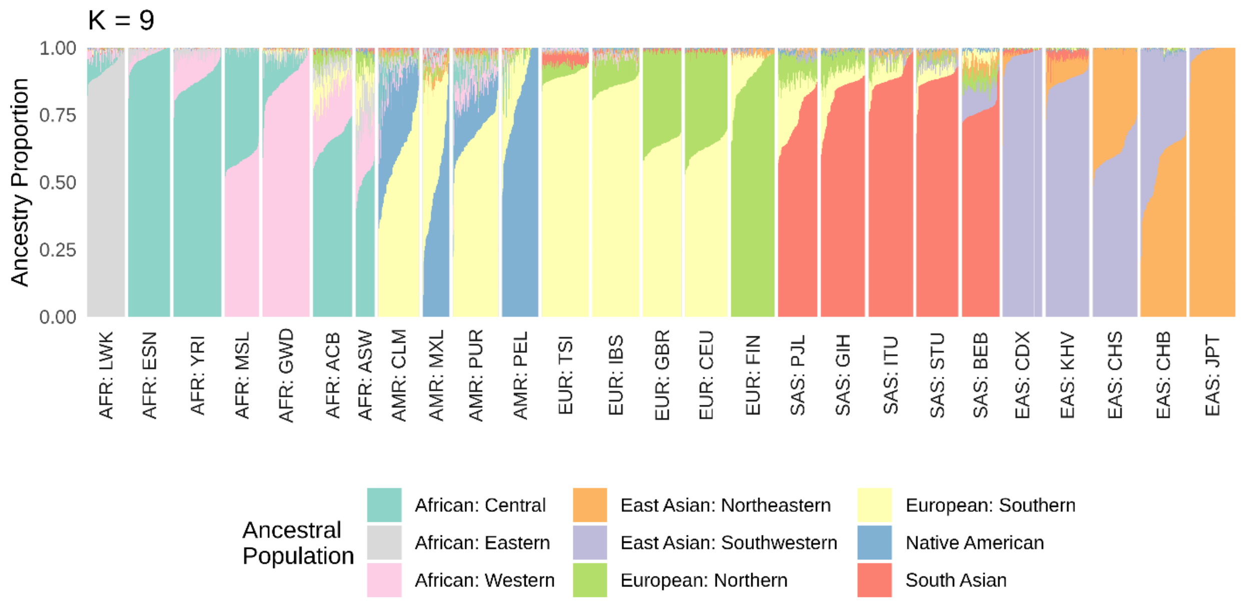

### Supplemental Tables

**Supplemental Table 1.** Demographic characteristics and comorbidities of first-degree relatives, by genetic risk group and age group

| | Negative FDRs (N = 58) | | | | VUS FDRs (N = 27) | | LP/P FDRs (N = 39) | |
|---|---|---|---|---|---|---|---|---|
| | *FDRs of probands with P/LP variants* (***reference***) (N = 28) | | *FDRs of probands without P/LP variants* (N = 30) | | | | | |
| **Characteristic** | **Below median age[a] (N = 14)** | **Above median age[a] (N = 14)** | **Below median age[a] (N = 14)** | **Above median age[a] (N = 16)** | **Below median age[a] (N = 13)** | **Above median age[a] (N = 14)** | **Below median age[a] (N = 21)** | **Above median age[a] (N = 18)** |
| **Demographics** | | | | | | | | |
| Age at echocardiogram, years, median (IQR) | 34.0 (28.0, 39.1) | 60.3 (54.9, 63.1) | 38.5 (30.1, 39.8) | 62.7 (55.3, 63.9) | 27.9 (19.7, 41.1) | 57.2 (49.4, 68.2) | 28.3 (25.4, 32.0) | 60.1 (51.9, 65.5) |
| Female, No. (%) | 9 (64.3) | 9 (64.3) | 9 (64.3) | 10 (62.5) | 8 (61.5) | 9 (64.3) | 14 (66.7) | 13 (72.2) |
| **Comorbidities, No. (%)** | | | | | | | | |
| Obesity (BMI ≥30 kg/m$^2$) | 6 (42.9) | 9 (64.3) | 7 (50.0) | 7 (43.8) | 2 (15.4) | 4 (28.6) | 4 (19.0) | 6 (33.3) |
| Hypertension | 0 (0.0) | 4 (28.6) | 0 (0.0) | 6 (37.5) | 0 (0.0) | 3 (21.4) | 1 (4.8) | 4 (22.2) |
| High Cholesterol | 0 (0.0) | 2 (14.3) | 0 (0.0) | 1 (6.3) | 0 (0.0) | 0 (0.0) | 0 (0.0) | 3 (16.7) |
| Diabetes | 0 (0.0) | 0 (0.0) | 0 (0.0) | 1 (6.3) | 0 (0.0) | 1 (7.1) | 0 (0.0) | 0 (0.0) |
| Tobacco use (ever), No. (%) | 3 (21.4) | 2 (14.3) | 5 (35.7) | 2 (12.5) | 4 (30.8) | 4 (28.6) | 6 (28.6) | 2 (11.1) |
| Years smoked, mean (SD) | 16.7 (5.8) [n=3] | 32.5 (10.6) [n=2] | 11.7 (7.0) [n=5] | 14.0 (5.7) [n=2] | 18.3 (14.4) [n=3] | 24.3 (13.1) [n=4] | 6.8 (4.8) [n=6] | 21.5 (19.1) [n=2] |
| Cigarettes per day, mean (SD) | 15.5 (13.4) [n=2] | 25.0 (7.1) [n=2] | 15.6 (15.1) [n=5] | 10.0 (14.1) [n=2] | 8.7 (10.3) [n=3] | 15.8 (5.1) [n=4] | 11.8 (9.7) [n=4] | 10.0 (-) [n=1] |
| Alcohol use (ever) and consume >=5 drinks in an occasion, No. (%) | 1 (7.1) | 1 (7.1) | 1 (7.1) | 1 (6.3) | 1 (7.7) | 1 (7.1) | 2 (9.5) | 2 (11.1) |
| Alcohol use frequency, No. (%) | [n=12] | [n=13] | [n=8] | [n=9] | [n=7] | [n=9] | | [n=14] |

| Characteristic | Negative FDRs (N = 58) | | | | VUS FDRs (N = 27) | | LP/P FDRs (N = 39) | |
|---|---|---|---|---|---|---|---|---|
| | *FDRs of probands with P/LP variants* (***reference***) (N = 28) | | *FDRs of probands without P/LP variants* (N = 30) | | | | | |
| | **Below median age[a] (N = 14)** | **Above median age[a] (N = 14)** | **Below median age[a] (N = 14)** | **Above median age[a] (N = 16)** | **Below median age[a] (N = 13)** | **Above median age[a] (N = 14)** | **Below median age[a] (N = 21)** | **Above median age[a] (N = 18)** |
| 1-3 times/month or <1 time/month | 3 (25.0) | 5 (38.5) | 4 (50.0) | 6 (66.7) | 5 (71.4) | 6 (66.7) | 2 (9.5) | 4 (28.6) |
| 1-3 times/week | 9 (75.0) | 5 (38.5) | 2 (25.0) | 2 (22.2) | 2 (28.6) | 2 (22.2) | 11 (52.4) | 5 (35.7) |
| 4-7 times/week | 0 (0.0) | 3 (23.1) | 2 (25.0) | 1 (11.1) | 0 (0.0) | 1 (11.1) | 8 (38.1) | 5 (35.7) |

Abbreviations: FDR = first-degree relative; LP = Likely pathogenic; LV = left ventricular; P = pathogenic; VUS = variant of uncertain significance.
[a] The median age of FDRs in the sample was 44.9 years.

**Supplemental Table 2.** Echocardiographic measurements of first-degree relatives, by genetic risk group and age group

| | **Negative FDRs (N = 58)** | | | | **VUS FDRs (N = 27)** | | **P/LP FDRs (N = 39)** | |
|---|---|---|---|---|---|---|---|---|
| | *FDRs of probands with P/LP variants* (***reference***) (N = 28) | | *FDRs of probands without P/LP variants* (N = 30) | | | | | |
| **Echocardiographic Measurement** | **Below median age[a] (N = 14)** | **Above median age[a] (N = 14)** | **Below median age[a] (N = 14)** | **Above median age[a] (N = 16)** | **Below median age[a] (N = 13)** | **Above median age[a] (N = 14)** | **Below median age[a] (N = 21)** | **Above median age[a] (N = 18)** |
| LV global longitudinal strain[b], %, mean (SD) | 20.8 (2.9) | 22.4 (1.8) [n=12] | 20.0 (2.0) | 21.2 (1.9) | 20.9 (2.4) [n=11] | 20.0 (2.0) | 20.4 (2.6) [n=18] | 18.8 (2.4) [n=17] |
| LV global longitudinal strain category[b], No. (%) | | [n=12] | | | [n=11] | | [n=18] | [n=17] |
| ≥18% (normal) | 11 (78.6) | 12 (100.0) | 11 (78.6) | 16 (100.0) | 10 (90.9) | 13 (92.9) | 15 (83.3) | 11 (64.7) |
| ≥16%, <18% (borderline) | 3 (21.4) | 0 (0.0) | 3 (21.4) | 0 (0.0) | 1 (9.1) | 1 (7.1) | 2 (11.1) | 4 (23.5) |
| <16% (abnormal) | 0 (0.0) | 0 (0.0) | 0 (0.0) | 0 (0.0) | 0 (0.0) | 0 (0.0) | 1 (5.6) | 2 (11.8) |
| LV longitudinal strain (A4C view) , %, mean (SD) | 20.9 (3.2) | 22.2 (2.2) | 19.4 (2.4) | 21.0 (2.6) | 21.4 (2.7) | 20.0 (1.9) | 20.4 (3.3) | 19.1 (2.9) |
| LV ejection fraction, %, mean (SD) | 60.4 (3.6) | 63.3 (3.7) | 59.9 (3.9) | 61.9 (3.1) | 61.0 (3.7) | 60.8 (3.5) | 59.1 (3.4) | 60.0 (3.5) |
| LV internal diameter at end diastole, z-score[c], mean (SD) | 0.1 (1.1) | -1.3 (1.5) | 0.5 (1.0) | -0.7 (1.4) | -0.4 (1.0) | -1.8 (2.4) | 0.0 (1.0) [n=20] | -0.7 (1.1) |
| LV internal diameter at end systole, z-score[c], mean (SD) | 0.9 (0.9) | -0.7 (1.0) | 1.4 (0.9) | 0.1 (1.6) | 0.4 (0.7) | -0.5 (1.3) | 1.0 (1.3) [n=20] | 0.6 (1.0) |
| Indexed LV end diastolic volume[d], ml/$m^2$, mean (SD) | 58.0 (8.0) | 48.0 (8.3) | 55.8 (9.6) | 45.6 (8.5) | 51.6 (11.1) | 48.8 (10.6) | 55.7 (8.8) | 46.7 (9.8) |
| Indexed LV end systolic volume[d], ml/$m^2$, mean (SD) | 22.9 (3.1) | 17.7 (3.8) | 22.3 (4.2) | 17.4 (3.7) | 20.0 (4.0) | 19.2 (4.8) | 22.8 (4.0) | 18.8 (4.7) |

| Echocardiographic Measurement | **Negative FDRs (N = 58)** | | | | **VUS FDRs (N = 27)** | | **P/LP FDRs (N = 39)** | |
|---|---|---|---|---|---|---|---|---|
| | *FDRs of probands with P/LP variants* (***reference***) (N = 28) | | *FDRs of probands without P/LP variants* (N = 30) | | | | | |
| | **Below median age[a] (N = 14)** | **Above median age[a] (N = 14)** | **Below median age[a] (N = 14)** | **Above median age[a] (N = 16)** | **Below median age[a] (N = 13)** | **Above median age[a] (N = 14)** | **Below median age[a] (N = 21)** | **Above median age[a] (N = 18)** |
| Posterior wall thickness at end diastole, mm, mean (SD) | 7.6 (1.4) | 8.6 (1.0) | 8.3 (1.4) | 8.9 (1.4) | 7.3 (1.3) | 8.5 (0.9) | 7.5 (1.3) [n=20] | 8.6 (1.1) |
| Septal wall thickness at end diastole, mm, mean (SD) | 8.4 (1.7) | 11.2 (2.5) | 8.7 (1.7) | 10.1 (1.4) | 7.9 (1.6) | 9.7 (1.5) | 7.8 (1.6) [n=20] | 9.6 (2.3) |

Abbreviations: FDR = first-degree relative; LP = Likely pathogenic; LV = left ventricular; P = pathogenic; VUS = variant of uncertain significance.

[a] The median age of FDRs in the sample was 44.9 years.

[b] LV global longitudinal strain could not be quantified in n = 8 FDRs who were missing one or more of the apical 2- or 3-chamber views.

[c] Calculated based on sex and height[35] for all study participants with heights of at least 152 cm (male) or 137 cm (female).

[d] Calculated using volume divided by Mosteller body surface area.

**Supplemental Table 3.** Covariate-adjusted estimated mean differences in echocardiographic measurements between positive FDRs (LP/P FDRs and VUS FDRs) and negative FDRs of probands without P/LP variants

| Measurement<br>Age group[a] | VUS FDRs | | P/LP FDRs | |
|---|---|---|---|---|
| | **Estimate (95%CI)[b]** | **P[b]** | **Estimate (95%CI)** | **P[b]** |
| LV global longitudinal strain, % | | | | |
| Below median | 0.1 (-3.0, 3.2) | 0.95 | -0.2 (-2.2, 1.7) | 0.82 |
| Above median | -1.8 (-3.1, -0.6) | 0.004 | -2.6 (-4.0, -1.2) | <0.001 |
| LV longitudinal strain (A4C view) , % | | | | |
| Below median | 1.5 (-1.1, 4.0) | 0.26 | 0.7 (-1.3, 2.7) | 0.50 |
| Above median | -1.6 (-3.2, -0.1) | 0.04 | -2.1 (-4.0, -0.3) | 0.03 |
| LV ejection fraction[d], % | | | | |
| Below median | 1.2 (-1.9, 4.3) | 0.46 | -0.8 (-3.6, 2.0) | 0.58 |
| Above median | -1.2 (-3.6, 1.3) | 0.35 | -2.1 (-5.0, 0.7) | 0.14 |
| LV internal diameter at end diastole, z-score[c] | | | | |
| Below median | -0.6 (-1.8, 0.5) | 0.29 | -0.5 (-1.2, 0.3) | 0.24 |
| Above median | -0.9 (-2.3, 0.4) | 0.18 | 0.0 (-1.0, 0.9) | 0.93 |
| LV internal diameter at end systole, z-score[c] | | | | |
| Below median | -0.7 (-1.5, 0.1) | 0.10 | -0.2 (-0.9, 0.5) | 0.63 |
| Above median | -0.4 (-1.4, 0.6) | 0.46 | 0.5 (-0.6, 1.5) | 0.37 |
| Indexed LV end diastolic volume, $ml/m^2$ | | | | |
| Below median | -3.7 (-10.9, 3.5) | 0.31 | 0.3 (-7.3, 7.8) | 0.94 |
| Above median | 3.3 (-2.9, 9.5) | 0.30 | 1.1 (-6.1, 8.3) | 0.77 |
| Indexed LV end systolic volume, $ml/m^2$ | | | | |
| Below median | -2.1 (-5.3, 1.1) | 0.19 | 0.6 (-2.8, 4.0) | 0.72 |
| Above median | 1.9 (-0.7, 4.5) | 0.15 | 1.5 (-1.8, 4.8) | 0.39 |

| Measurement<br>Age group[a] | VUS FDRs | | P/LP FDRs | |
|---|---|---|---|---|
| | Estimate (95%CI)[b] | P[b] | Estimate (95%CI) | P[b] |
| Posterior wall thickness at end diastole, mm | | | | |
| Below median | -0.9 (-2.1, 0.4) | 0.17 | -0.5 (-1.7, 0.7) | 0.42 |
| Above median | -0.2 (-1.1, 0.7) | 0.62 | -0.1 (-1.4, 1.1) | 0.82 |
| Septal wall thickness at end diastole, mm | | | | |
| Below median | -0.9 (-2.4, 0.5) | 0.20 | -0.8 (-2.3, 0.7) | 0.29 |
| Above median | -0.2 (-1.7, 1.2) | 0.73 | -0.4 (-2.0, 1.1) | 0.58 |

Abbreviations: A4C = apical 4-chamber; DCM = dilated cardiomyopathy; FDR = first-degree relative; P = pathogenic; LP = Likely pathogenic; LV = left ventricular; VUS = variant of uncertain significance.

[a] Below or above the median age of FDRs in the sample (44.9 years).

[b] The mean of each echocardiographic measurement was modeled as a function of genetic risk group within two age groups (below and above the median age in the sample) using a single linear mixed model with an interaction. This model specification was chosen a priori to reflect expected growth in the differences between genetic risk groups with age under a threshold model for DCM development that accounts for age-dependent penetrance. For all measurements other than sex-specific internal diameter z-scores, sex and its interaction with age group were included as covariates. For LV global longitudinal strain and LV longitudinal strain (A4C view), height, weight, and image quality rating (≤2 vs. >2) were also included. Heterogeneity between clinical sites and intrafamilial correlation were modeled by including independent normal random effects for proband enrollment site and family within site. Covariate-adjusted estimated differences in means between each positive FDR group (LP/P FDRs and VUS FDRs) and negative FDRs of probands without P/LP variants, their 95% confidence intervals, and two-sided Wald p-values for the null of no difference were obtained from this model using Morel-Bokossa-Neerchal bias-corrected empirical standard errors and the standard normal distribution. Estimated marginal means for each genetic risk group derived from this model are shown in Figure 3 (FDRs above the median age) and Supplemental Figure 1 (FDRs below the median age). Table 2 shows the number of FDRs contributing to each measurement's model by genetic risk group.

[c] The final model excluded the site random effect because convergence occurred on the boundary constraint with a zero variance component when it was included. Morel-Bokossa-Neerchal bias-corrected empirical standard errors with sites as independent units were still used.

[d] The final model excluded the site and family-within-site random effects because convergence occurred on the boundary constraint with zero variance components when they were included. Morel-Bokossa-Neerchal bias-corrected empirical standard errors with sites as independent units were still used.

26. Richards S, Aziz N, Bale S*, et al.* Standards and guidelines for the interpretation of sequence variants: a joint consensus recommendation of the American College of Medical

Genetics and Genomics and the Association for Molecular Pathology. *Genet Med* 2015;**17**:405-424. doi: 10.1038/gim.2015.30
